# Adult Scoliosis and Exercise: A Survey Instrument Pilot Study

**DOI:** 10.1101/2025.04.11.25325623

**Authors:** Christine Whitmarsh

## Abstract

The relationship between scoliosis pain and exercise is described in the literature as inconsistent and highly subjective to each patient. Researchers cite a lack of understanding about the true nature and etiology of scoliosis pain, often referring to it as a “structural deformity (only).” This survey instrument pilot study aims to better understand scoliosis pain, filling in the gaps left by existing scales on scoliosis and chronic pain. The scale also aims to assess experiences and attitudes about utilizing exercise for pain relief. The 18-item survey instrument was distributed to a sample (n=95) of individuals who self-reported having adult scoliosis for 10 years or longer, who experience scoliosis pain, and who exercise. Reliability and exploratory factor analyses (EFA) were run on several scale subsets, with pain-related subsets and items showing higher reliability than exercise items. The most compelling result was the 3-factor solution that emerged to describe possible latent pain variables - mobility, neurological, and stress effects. This small, initial pilot study is the initial step in a planned long-term research mission to help establish evidence-based recommendations that can potentially help a large population of adults with scoliosis who experience long-term, chronic pain and disability. Additional planned research includes a focus group as a qualitative means of obtaining richer data on types, frequency, dosage, and intensity of exercise found to be most helpful in managing the chronic pain and disability of adult scoliosis. A mixed methods study is also planned to aid in further quantifying the impacts of exercise on scoliosis pain, with a qualitative aspect to deepen the quantitative data obtained.

## Introduction

In Ancient Greece, Hippocrates (460-370 BC) first introduced the term “scoliosis” and developed his ideas about the spinal disease through his writings, including on topics like axial traction and trans-abdominal correction as potential corrective strategies. Nearly 500 years later, Galen built on Hippocrates’ work, exploring more in-depth the causes and etiology of scoliosis. Galen’s work would influence spinal medicine for over 1500 years. (Vasiliadis, et al., 2009).

Scoliosis is defined as a lateral spinal curvature greater than 10 degrees. While the exact cause of the disorder is unknown, possible reasons include genetic factors, biomechanical, neurological, or musculoskeletal issues, or growth and development factors (Chen, et al., 2024; Chen, et al., 2023). There is a 97% correlation in families where scoliosis is already present, indicating that at least one main gene is inherited (Chen, et al., 2024; Chen, et al., 2023). 2-4% of children will develop adolescent idiopathic scoliosis (AIS), the most common type of scoliosis (Chen, et al., 2024; Chen, et al., 2023).

In addition to its defining lateral curves, other signs of AIS include uneven leg length, uneven shoulders, and other postural issues. Severe cases of scoliosis can result in pain, anxiety, depression, fear, respiratory difficulties, and impaired cardiopulmonary function (Chen, et al., 2024; Chen, et al., 2023). 10-15% of adolescents with mild AIS are at risk for worsening curve (Li, et al., 2021). Girls are nearly 6 times more likely to show more symptoms than boys (An, et al., 2023). AIS is the third most common health issue in adolescents after obesity and myopia (Chen, et al., 2024; Chen, et al., 2023).

Curves of 25 or less degrees in adolescents or 40 or less degrees in adults typically warrant only observation, 25-45 degrees, bracing, and when scoliosis curves are 45 degrees or greater, surgery is typically considered since these curves have a high rate of worsening even into adulthood (An, et al., 2023).

### Scoliosis Pain

Pain typically shows up as a side effect of scoliosis when curvatures reach 30 degrees or higher by adulthood (An, et al., 2023; Li, et al., 2021). Some research shows that the more severe the curvature (measured by the Cobb Angle and spinal rotation), the higher the pain intensity and lower quality of life (An, et al., 2023; Li, et al., 2021). With degenerative (versus AIS) pain can set in at even smaller curvatures (Zaina, et al., 2023). One multivariate analysis showed patients with thoracic curves had significantly increased pain in contrast to those with lumbar curves (An, et al., 2023).

There are differences in pain presentation and effects between adults and adolescents with scoliosis. One study noted that while factors such as curvature and spinal deformity, both primarily measured via x rays, mandate treatment in younger patients, for adult scoliosis patients, treatment is dictated by pain and disability, with the additional observation that level of disability cannot be predicted solely by x rays (Bess, et al., 2009). Scoliosis related groin/pelvic pain and its related radiation to the legs seems most associated with lower back curvatures, which combined, can increase lower back pain intensity for adults with scoliosis (Gremeaux, et al. 2008).

Up to 37.6% of people 60 years old and old, report having back pain. But studies show people with scoliosis are more likely to have more severe and a longer duration of pain than those with back pain but no scoliosis. Scoliosis pain is more asymmetric than regular back pain and can radiate to one or both legs. It tends to increase with prolonged standing, and reduce when lying down. Those with back pain from scoliosis usually need to change position more frequently as well (Zaina, et al., 2023).

Yet, even in light of this research, the relationship between scoliosis and pain remains inconclusive. Evidence shows that while adults with scoliosis frequently report pain, whether the pain is related to their spinal curvature, is not always clear (Zaina, et al., 2023). One piece of literature found a 40% lifetime prevalence of back pain in adolescents in ages 9-18 (An, et al., 2023). Another team of researchers reflected on how AIS was once considered a painless condition, until a conglomerate of studies showed pain occurring in a “significant proportion” of scoliosis patients (Djurasovic, et al., 2018).

Dr. Arnold Y. L. Wong made several important discoveries regarding scoliosis pain and clinical recommendations in the collaborative 2019 work “How Common Is Back Pain and What Biopsychosocial Factors Are Associated With Back Pain in Patients With Adolescent Idiopathic Scoliosis?” as recounted to Dr. Seth S. Leopold (Leopold, 2019). Dr. Wong pointed out how both clinicians and patients alike, often focus more on variables like the magnitude of the physical deformity and curve progression than actual symptoms, such as pain associated with the disorder (Leopold, 2019). Wong went on to state that “most clinicians” see back pain in scoliosis patients as something that does not occur until 30 or 40 years following initial diagnosis, calling it surprising, in his work, to discover the prevalence of back pain in teenagers with scoliosis with nearly 9% of AIS patients reporting back pain persisting for more than 3 months, therefore clinically qualifying as chronic pain (Leopold, 2019).

There is also an overall lack of correlation between spinal curvature and severity of pain with “limited and contradictory evidence” regarding the potential benefits of treatments designed to reduce curves and pain reduction (An, et al., 2023).

Researchers have cited a need to better understand the nature of back pain in those with scoliosis and determine more definitively whether scoliosis is at the root of the pain. “Despite the high prevalence of adult scoliosis (AS), there is an important gap in the literature with limited evidence reporting the effect of exercise on back pain in adults with scoliosis. This review suggests further experimental research is needed and formulates research recommendations” (Alanazi, et al., 2018, p. 1). With a clearer understanding of scoliosis pain, it is felt that more specific treatment strategies can be implemented (Zaina, et al., 2023). Another study concurs that while “Back pain is an important consideration in treatment,” (An, et al., 2023, p. 127), “Back pain in AIS is common but remains difficult to predict and treat.” (An, et al., 2023, p. 126).

### Modifiers of Scoliosis Back Pain

There have also been found to be primarily psychological modifiers of back pain experienced by those with scoliosis, including patient catastrophizing (assuming the very worst about a situation including that it will have no end), anxiety, and mood (An, et al., 2023). In fact, when patients with AIS were coached on viewing scoliosis as a manageable condition, rather than a serious disease that could take over every aspect of their lives, and taught to manage things like depression and fatigue while reigning in the tendency to catastrophize, it led to improved agency over their pain management and an overall improved reported quality of life (An, et al., 2023).

### Surgery as a Corrective Mechanism

The method of treatment for scoliosis depends on the degree of curvature(s), age of the patient, and skeletal maturity. Conservative forms of treatment like brace-wearing or exercise are indicated to slow or stabilize curve progression, where surgery is indicated for the most severe cases with no other viable options, since surgery causes severe tissue trauma and serious postoperative pain (Chen, et al., 2024).

Unfortunately, surgery does not assure a pain free future for those with scoliosis. In one metaanalysis, 3 of 7 studies showed that, based on the SRS-24 scoliosis pain/functionality instrument, at 24 months post op, pain scores became worse than preoperative ones and after 60 months, 2 of 3 studies showed worse pain compared to pre surgical pain levels (An, et al., 2023).

In 2023, a comparative cohort study looking at back pain and quality of life 10 years after posterior spinal fusion surgery, showed pain score improvement 2 years post op but at 10 years, those gains were largely wiped out as pain score returned to preoperative levels (Ahonen, et al., 2023). The sole lasting gains from the surgery, using the SRS-24 instrument were in the area of self image (Ahonen, et al., 2023).

“Several studies suggest pain may improve initially but may recur over time even to baseline levels.” (An, et al., 2023, p. 136). Scoliosis patients who underwent surgery, in addition to experiencing a recurrence of preoperative pain, also reported a decline in overall functionality following surgery (An, et al., 2023). Similar to the psychological moderators of the pain itself, in the same studies, preoperative pain scores moderated by anxiety, mood, and overall mental health, accurately predicted postoperative pain scores (An, et al., 2023).

### Exercise and Scoliosis Pain

The predominance of research on the subject covered in this review indicates that exercise is utilized in the treatment of scoliosis to control the rate of curve progression, create skeletomuscular balance and improve aesthetics, reduce surgical rates, and improve overall quality of life (An, et al., 2023; Gur, et al., 2017). Exercise is, however, noted to be an effective method of pain relief (Gur, et al., 2017). The best results occurred after long-term exercise programs, with one example including a 35 month long “scoliosis intensive” program (Gur, et al., 2017).

Not all exercises are created equal, especially when it comes to the specialized needs and structural abnormalities of scoliosis. Traditional exercises, meaning exercises other than core strength and/or stability, pilates, or scoliosis specific exercises, have not been shown to significantly improve pain levels or quality of life (Gur, et al., 2017). In a meta-analysis of studies examining scoliosis and exercise interventions, scoliosis specific exercises were shown to be more effective than traditional exercise in stabilizing and/or reducing degree of curvature, although there were no statistically significant differences in pain levels (Gur, et al., 2017).

In regards to curve reduction, the most effective forms of exercise for scoliosis were, in order: yoga, core strength/stability, PSSE (physiotherapeutic scoliosis specific exercise), Schroth training (a form of PSSE customized to severity and type of curve), and sling exercises (Chen, et al., 2023). Researchers concluded overall, in this analysis, that exercise has the capacity to significantly improve the curvature measurements of AIS and more so than via other types of scoliosis therapy (Chen, et al., 2023).

Taking a closer look at some of these exercise types and results achieved, one randomized clinical trial of 110 patients showed better results with PSSE than conventional physical therapy (PT) in patients 10 years of age and older with small curves 10-25 degrees (Marti, et al., 2015). It has also been shown that PSSE is prescribed less frequently than PT for pain relief (3%) than for improving aesthetics (62%), with the most common type of PSSE being Schroth training (57%) (Marti, et al., 2015). Compared to PT, Schroth exercises were more effective in improving patients’ stated goals, including postural balance, aesthetics, stability of curvature (reduced Cobb angle), and/or pain relief (Chen, et al., 2024).

Core stability and strength exercises have produced promising research evidence in treating scoliosis pain. Core stabilization exercises when combined with traditional exercise have been shown to be more effective than traditional exercise alone, in reducing scoliosis pain and accompanying spinal rotation (Gur, et al., 2017), along with improving Cobb angles and quality of life (Li, et al., 2021). Core training has also improved quality of life in patients with generalized lower back pain, however, comparable studies of scoliosis are lacking (Gur, et al., 2017), particularly in regards to pain relief for people with scoliosis, as measured by the SRS-22 survey instrument (Li, et al., 2021).

Once again, we find differences in adolescent (AIS) and adult scoliosis (AS), this time in the rationale to treat with exercise, where for AIS, as previously stated, the primary motivator is usually curve reduction while for adults where that is less possible due primarily to the effects of aging, the rationale for exercise becomes decreased pain and increased functional mobility. In a series of physical therapy case studies, 3 adults with adult scoliosis, aged 31, 48, and 68 years old, were treated with a combination of scoliosis-specific exercises (PSSE) and traditional physical therapy. All three patients reported decreased pain (using the traditional VAS pain scale) and improved functional mobility, along with increased ability to self-manage their pain symptoms (Roberts, et al., 2023).

### Adult Scoliosis (AS) and Exercise

A 2018 systematic review of literature on scoliosis back pain and exercise, initially found 630 possible articles for review, saw their batch weaned down to 98 after screening full study text, but of those 98, only 1 single study met the researchers’ review criteria, mostly due to study design and/or patient population. The one remaining study, a randomized controlled trial of 130 adult patients with idiopathic scoliosis found only limited evidence that 20 weeks of a combination of various (primarily core) stabilization exercises improved pain, disability, and quality of life in study participants (Alanazi, et al., 2018). The conclusion of this systematic review was that, “Despite the high prevalence of AS, there is an important gap in the literature with limited evidence reporting the effect of exercise on back pain in adults with scoliosis” (Alanazi, et al., 2018, p. 647).

One correlational study investigated the effects of two different, targeted methods of exercise on 62 adult scoliosis patients (mean age 31 years old) divided into experimental and control groups, over a 6 month period of time (Nikolov, et al., 2017). The experimental group received traditional physical therapy, as well as specific spinal mobilization and postural therapy techniques (Nikolov, et al., 2017). Following the course of treatment, patients in the experimental group reported an average of 2.75 reduction in pain levels using the visual analogue scale (VAS), with the control group, which received only traditional physical therapy, reporting a 1.88 reduction in pain (Nikolov, et al., 2017). No patients reported an increase in pain, and muscle strength in the trunk also increased in both groups (Nikolov, et al., 2017). “The combined use of spinal mobilization and postural therapy appeared to significantly reduce the levels of pain and increase trunk muscle strength in all 62 subjects” (Nikolov, et al., 2017, p. 33).

Another finding of the extended literature review, was that there appears to be no lack of case studies, or other small studies showing a potential inverse relationship between exercise and pain in adults with scoliosis. Blum (2002) performed a case study on a 39 year old woman with scoliosis who was approximately 20 years post spinal fusion surgery, but was experiencing worsening severe low back pain. A prescribed course of Pilates exercises (subjectively) helped stabilize the patient’s pain, allowing her to gradually increase physical activity (Blum, 2002).

A 37 year old patient with adult scoliosis underwent a Schroth scoliosis exercise program along with part-time back bracing and 16 months following completion of the program, reported being free of what had been daily low back pain (Weiss, et al., 2016). Objectively, the patient’s spinal curvature, measured by Cobb angle, and angle of trunk rotation had also improved (Weiss, et al., 2016).

Roberts et al. (2023) performed a case study series of three patients, aged 31, 48, and 68, with adult scoliosis and hip pain, implementing PSSE (scoliosis-specific physio type exercises) into their existing physical therapy treatments. At the completion of the combined therapy, all three patients reported decreased pain, as measured by the Visual Analog Scale, as well as increased functional mobility in their activities of daily living (Roberts, et al., 2023).

These are presumably an extremely small representation of the case studies and other small sample, qualitative, and mixed methods oriented explorations of the relationship between adult scoliosis pain and exercise as a potential method of pain relief. There appears to be a scant amount of experimental research with large sample sizes and methodological rigor, to provide more quantifiable, high value, and, ideally, generalizable, research evidence on the subject of adult scoliosis pain and exercise.

Another example of issues with generalizability is found in the case of Nikolov et al. (2017), in what appeared to be ultra specific exercise techniques that were applied as the treatment variable, raising the question of external validity via generalizability of the treatment variable to the larger population of adults with scoliosis. When it comes to the research potential of adult scoliosis and exercise as a pain management strategy, the runway of possibility appears to be long.

### Types of Exercise and Adult Scoliosis

#### I. Scoliosis-Specific Exercises (PSSE, Schroth, SEAS): Based on the totality of evidence reviewed, scoliosis-specific exercise appears to be more effective than “traditional” exercise methods as pain management for adults with scoliosis

○ Compared to PT, Schroth exercises were more effective in improving patients’ stated goals, whether postural balance, aesthetics, stability of curvature (reduced Cobb angle), or pain relief (Chen, et al., 2024).
○ Exercise is considered to be a conservative scoliosis treatment. One set of studies reported in a 2019 synthesis placed the success rate of conservative forms of treatment at just 27% (Ng, et al., 2019).
○ In the same 2019 synthesis, physiotherapeutic scoliosis-specific exercises (PSSE) when combined with several other forms of conservative therapies resulted in lower pain and disability ratings in adult scoliosis patients with thoracolumbar and lumbar curves (middle/low back and low back). Multiple studies supported this, with adult scoliosis patients reporting less pain after undergoing a PSSE condition (Ng, et al., 2019).
○ The same synthesis wrote about a group of young women with varying degrees and locations of curvatures who took part in a series of case studies, also reported a significant drop in lower back pain after 1 month of taking part in PSSE (Ng, et al., 2019).
○ A 2015 study focused on the loss of sagittal balance as a key factor in increased pain and disability in adult scoliosis. The study concluded that PSSEs might create physical compensation that can decrease pain and disability, thus increasing overall quality of life for those with scoliosis (Negrini, et al., 2015).

#### II. Pilates/Core Exercises: In the category of non-scoliosis specific exercises, however, pilates/core exercises have shown some very promising results including in a handful of RCTs

○ Core stabilization exercises when combined with traditional exercise have been shown to be more effective than traditional exercise alone, in reducing scoliosis pain and accompanying spinal rotation (Gur, et al., 2017)
○ “The combined use of spinal mobilization and postural therapy appeared to significantly reduce the levels of pain and increase trunk muscle strength in all 62 subjects” (Nikolov, et al., 2017, p. 33).
○ Moving on to Pilates, another type of exercise that some research has shown to be an effective pain management strategy for those with scoliosis. A 2012 experimental study divided a sample of 31 female students into a control group (no Pilates) and experimental group (Pilates). The pain scale used, as measured by a dependent t test, showed a significant reduction in pain and increased flexibility in the experimental group (Araújo, et al., 2012).
○ Most recently, a 2022 meta-analysis (118 trials, n=9710) found that the experimental groups prevailed, and that all of the physical exercises included in the intervention resulted in improved pain and disability in participants. The top 3 for pain reduction were Pilates, mind-body, and core exercises. Pilates, strength, and core exercise, were most effective in reducing disability. The analysis also found that the most beneficial interventions included a minimum of 1-2 sessions per week of Pilates or strength exercises for less than 60 minutes, for a range of 3-9 weeks. A SUCRA statistical analysis, which ranks interventions by how likely they are to be the best option, ranked Pilates as the most likely to reduce pain (93%) and lessen disability (98%) (Fernández-Rodríguez, et al., 2022).
○ Of all the research reviewed in response to the question, “if exercise does help scoliosis pain - which type(s) work best?”the one above, featuring Pilates and core training, might be one of the most valuable future paths forward to explore. This will require further detailed investigation of this meta-analysis, using CRC strategies.
○ (Negatives) A 2016 experimental study (n=130, curves <35 degrees) matched up various cognitive, non-exercise forms of rehabilitation (experimental group) against general forms of physiotherapy, stretching and core muscle strengthening (control group) over a 20-week period. The DVs included multiple pain and disability measurement scales including one specific to scoliosis. The experimental group (non-exercise) prevailed, showing improvements in pain and disability levels among participants that lasted for one year or longer (Monticone, et al., 2016).

#### III. Yoga: The evidence in this area was not nearly as promising for the other types of exercise explored, but still worthy of further exploration in terms of how yoga might assist with pain management for scoliosis

○ A broader 2011 experimental study, looked at the impact of a 12 week yoga intervention on 24 adults with chronic back pain, scoliosis patients included. The intervention included weekly group yoga classes as well as home practice, in several aspects of yoga. Using a self-report questionnaire as the method of measurement, repeated measures (baseline, 12 and 24 weeks) and one-sample t-tests showed statistically significant improvements in a variety of areas that included my areas of interest - disability, physical functioning, and physical pain. The highest ratings came at the 24 week measurements. The quantitative tests were triangulated with qualitative reporting by the participants that included reduced pain, more flexibility, and overall increased back strength. The main thing that might have improved the strength of these results would be if the sample had been made up exclusively of adult scoliosis patients (Schultz, et al., 2011).

#### IV. In Summary

○ The predominance of research reviewed so far, indicates that the main usage of exercise in the treatment of scoliosis is to control the rate of curve progression, create skeletomuscular balance and improve aesthetics, reduce surgical rates, and improve overall quality of life (An, et al., 2023; Gur, et al., 2017)
○ Exercise is, however, noted to be an effective method of pain relief (Gur, et al., 2017).
○ The best results occurred after long-term exercise programs, with one example including a 35 months “scoliosis intensive” program (Gur, et al., 2017).

### Limitations in Prescribing Exercise for Scoliosis Pain

Historically in orthopedic medicine, largely due to lack of convincing evidence, surgeons in the U.S. have questioned the efficacy of prescribing scoliosis specific exercise (like PSSE or PT) as treatment options for their scoliotic patients, but Marti (2015), notes that those attitudes appear to be evolving.

In one survey of scoliosis providers, Standard PT was used primarily to treat smaller curves (48%) or post operatively (37%) and in only 25% of the cases for pain relief (Marti, et al., 2015). PSSE was used to decrease scoliosis pain 2% of the time (Marti, et al., 2015). 63% of scoliosis clinicians prescribed neither PT nor PSSE, 11% prescribed both, 15% PT only, and 11% PSSE only. Reasons cited by the scoliosis providers surveyed for not prescribing exercise to their patients included: insufficient research (73%), no perceived value (53%), and no patient interest (6%) (Marti, et al., 2015). One conclusion of the survey was, “Despite an increased patient awareness of nonsurgical treatments for AIS, the majority of surgeons do not prescribe PT/PSSE.” (Marti, et al., 2015, p. 5). However, most of the surveyed scoliosis providers asserted that in the face of more research evidence that exercise works, whether PT, PSSE or other, they might consider prescribing it more often (Marti, et al., 2015).

A separate 2019 survey of Scoliosis Research Society members, entirely spine surgeons, examined attitudes toward prescribing traditional physical therapy (PT) and PSSE scoliosis exercises for scoliosis (Steinmetz, et al., 2020). 64% of the surgeons prescribed PT to patients,, 40.3% did so for persistent pain, 34.3% for balance/gait issues in their patients, 25.4% prescribed PT in response to difficulty with daily activities, 74% of the surgeons were familiar with PSSE scoliosis exercises, and 66% felt comfortable prescribing PSSE exercises postoperatively (Steinmetz, et al., 2020). As for reasons for not prescribing physical therapy to their scoliosis patients, 50% of the surgeons cited lack of perceived value, 31.3% cited lack of evidence supporting its benefits, and 18.8% pointed to a lack of appropriate trained therapists (Steinmetz, et al., 2020).

### Literature Review Limitations

The demographics of the Gur (2017) study seemed reflective of the vast majority of studies cited in this literature review, where participants had adolescent idiopathic scoliosis, were approximately 10-16 years of age, with no stated comorbidities, most wore back braces already, and those who had undergone surgical correction were excluded. This combination of sample demographics along with the preponderance of methodology evidence presented in this review, would appear to indicate that a primary goal of research exploring the relationship between scoliosis and exercise is curve stabilization/correction (via Cobb Angles) more so than exercise as a form of pain management in scoliosis patients. There appear to be few studies looking at connections between exercise and pain and/or quality of life for those with scoliosis (Gur, et al., 2017).

### Conclusion of Literature Review

Far from a “crooked spine,” scoliosis is a complex disorder involving curvature, rotation, and asymmetry of the spine along with associated muscular and skeletomuscular issues affecting sometimes the entire torso of the afflicted person. In addition to the physical issues and limitations associated with scoliosis, there are also psychological moderators often present, including anxiety, depression, catastrophizing, and mood.

Recent research has highlighted the potential value and promise of exercise, especially scoliosis-specific forms like PT, PSSE, and core strengthening and stability, as methods of stabilizing curve progression and improving overall quality of life. Exercise as a method of pain management for those with scoliosis, however, seems less researched and therefore less understood by scoliosis clinicians. This literature review underscores an opportunity for more comprehensive research on the role of exercise as a pain management strategy for those with scoliosis.

The research also highlights a need to expand research into more diverse scoliosis patient populations, beyond adolescents with adolescent idiopathic scoliosis and also the need for research that is inclusive of those who have undergone corrective scoliosis surgery and are still in need of long-term pain management strategies.

### Pilot Study Construct of Interest

The goal of this pilot study was to develop a survey instrument that gathers subjective data about the experience of living with chronic scoliosis pain as well as the impact of exercise on pain levels and functionality in adults with chronic pain from scoliosis.

### Overview of Existing Survey Instruments on This Topic

#### Scoliosis

##### SRS-22r

This scale appears to be the benchmark among orthopedic specialists, of measuring the impacts and outcomes of scoliosis in patients over 10 years of age. The 22-item questionnaire covers 5 domains of scoliosis including - function/activity, pain, mental health, self image, and management satisfaction. This scale has been shown to have good/excellent test-retest reliability and internal consistency (Gardner, et al. 2021; Gur, et al. 2017).

#### Chronic Pain

##### Oswestry Disability Index (ODI)

The ODI is a questionnaire for individuals with acute or chronic low back pain. It subjectively scores the level of disability in relation to activities of daily living (ADL), therefore making it an effective instrument for those suffering from severe levels of disability (Physiopedia, 2024).

##### Visual Analog Scale for Pain (VAS Pain)

One of the most common clinical and research measures to assess pain, patients pinpoint the intensity of their pain level on a 0-10 continuum (Price, et al., 1983).

##### Numeric Rating Scale for Pain (NRS Pain)

The NRS is similar to VAS except rather than having survey respondents pinpoint their level of pain with precision on a spectrum, the NRS uses numbers for rating pain intensity.

##### Measure of Intermittent and Constant Osteoarthritis Pain (ICOAP)

This scale is specifically designed to measure intermittent or constant pain associated with osteoarthritis.

##### Brief Pain Inventory (BPI)

While initially designed to measure cancer pain, the BPI has since been expanded to assess several chronic pain disorders as well. This scale assesses pain intensity and how much it disrupts daily functioning.

##### Chronic Pain Grade Scale (CPGS)

Chronic Pain Grade Scale/Questionnaire (CPGS) is a global scale that assesses the overall severity of chronic hip pain based on the intensity of the pain and disability related to the pain (Papaioannou et al., 2018). The CPGS “provides a categorical grading scheme and numerical self-rating scores for pain intensity and disability, allowing for qualitative changes in chronic pain over time” (Papaioannou et al., 2018, p. 38).

#### SEE-SV

This scale measures exercise as an effective chronic pain treatment, however with a third factor of self-efficacy either mediating or moderating (further exploration is required to determine which) the resulting treatment outcomes (Dahlbäck, et al., 2023).

#### McGill Pain Questionnaire

This scale converts patients’ subjective pain descriptors into “quantitative measures of clinical pain that can be treated statistically.” (Melzack, 1975).

#### SF-36

The SF-36 is a short-form health survey addressing eight aspects of health, with chronic pain appearing to be an implied causative factor for the areas, at least one of which addresses exercise (“limitations in physical activities”) (Ware, 1992).

### WHYMPI

The West Haven-Yale Multidimensional Pain Inventory (WHYMPI) is made up of 12 different scales that look at, among other things, how chronic pain impacts participants’ lives including its effect on daily activities (presumably exercise could be included in this) (Kern, et al., 1985).

### Potential Value of Existing Instruments

The previous scales offer prospective value and ideas that could be applied to this new instrument (which I have since tentatively named the “SE-18”), which would not be possible without the prior research contributions.

#### SRS-22

As what would appear to be the existing benchmark measure of scoliosis pain and functionality, it would be a mistake to overlook the SRS-22’s (and SRS-22R’s) value to adults with scoliosis. The shorter, more targeted SRS-22 has been noted to be reliable, with reproducibility, internal consistency, and concurrent validity as compared to the SF-36 scale, which is a broader assessment of overall health (Asher, et al., 2003). As a critique, while the SRS-22 covers many aspects of scoliosis, it only directly addresses the nature of scoliosis pain in a few questions.

#### Oswestry Disability Index (ODI)

As a scale dedicated to scoring disability in those with chronic back pain, the ODI’s language surrounding these topics will be studied and appropriately modeled.

#### Visual Analog Scale for Pain (VAS Pain)

Also known as the classic “0-10 pain scale” in clinical settings, the 0-10 continuum approach for scoring pain continues to be the unofficial flagship of pain assessment and will undoubtedly be utilized in many of the SE-18 pilot instrument items.

#### Numeric Rating Scale for Pain (NRS Pain)

The NRS is appealing due to its use of a spectrum for more specific response options to gauge intensity of pain.

#### Measure of Intermittent and Constant Osteoarthritis Pain (ICOAP)

Valuable here, is how the ICOAP distinguishes between intermittent and constant pain within a chronic pain condition assessment.

#### Brief Pain Inventory (BPI)

The BPI features a useful intersection of pain intensity and disruption of daily functioning.

#### Chronic Pain Grade Scale (CPGS)

Having already studied and worked with a CPGS dataset for a separate project, the value to be gleaned here is in this scale’s thorough approach to assessing the impact of pain on functioning, from day-to-day to various periods of time up to 6 months.

#### SEE-SV

Closely related to the SE-18 construct, the value of the SEE-SV might be how to effectively assess the impact of exercise as a treatment for chronic pain (including scoliosis).

#### McGill Pain Questionnaire

This instrument will be intriguing to explore in that it converts the qualitative to quantitative, something extremely relevant to the SE-18 goals, as well as planned, longer term mixed methods objectives in this work.

#### SF-36

How this instrument addresses chronic pain as a piece of the larger subject of overall health, could be interesting in regards to the item writing approach for the SE-18. The SF-36 was used in at least one piece of literature (Schwab, et al., 2003) to assess scoliosis pain in adults although that is not the primary indicated use of the scale. That study showed that adult scoliosis (the focus of the SE-18) has a negative impact on the perception of one’s overall state of health (Schwab, et al., 2003).

From a critical standpoint, only 2 questions in the SF-36 directly address pain. This scale appears to primarily assess pain more indirectly, through questions related to outcomes and collateral effects of living with pain (similar to how many of the scales covered here assess pain). The SF-36 showed evidence of validity and reliability when assessing populations with variable extents of ill health (Jenkinson, et al., 1994).

#### WHYMPI

There could be a collective takeaway from a review of this instrument regarding how to best measure the impact of chronic pain on functioning in scoliosis patients.

### Need for A Better Pain and Function Scale for Adults with Scoliosis

> *“It is our conclusion that adult scoliosis is becoming a medical condition of significant impact, affecting the fastest growing section of our society to a previously unrecognized degree.”*

> (Schwab, et al., 2003)

None of the chronic pain scales reviewed specifically addressed scoliosis pain. The scales in question measured either generalized chronic pain or other type-specific chronic pain such as hip or knee pain. And while there are numerous scales measuring chronic pain, there appear also to be a lack of scales that specifically measure the relationship between exercise and chronic pain management.

The sole scale specifically measuring pain and overall quality of life for individuals with scoliosis is the SRS-22 (and 22r). However, even this scoliosis-specific scale, as well as the other scales reviewed, tries to cover many different aspects of chronic pain management, including mental health elements such as self-efficacy, catastrophizing, self-image, anxiety, and depression, thus relegating exercise/physical movement to a secondary conversation rather than the primary one. While the majority of the pain scales in question, address intensity and frequency of pain, catastrophizing, Quality of Life (QOL) and Activities of Daily Living (ADL) - that is to say outcomes of pain and how pain ultimately impacts the individual’s life - most of the scales failed to address quality or location of pain.

The arguable exception to this assessment was the McGill and Brief Pain Inventory (BPI) scales. The McGill scale uses word selection charts to assist individuals describing the quality of their pain in great detail. The BPI provides a list of pain quality words with dichotomous yes/no response options. The BPI also offers a graphic of a human body where individuals could circle the locations of their pain. The SE-18 seeks to model the McGill word selection style in its item writing.

However, once again reinforcing the point that none of the scales studied, including the scoliosis oriented SRS-22, included items specifically addressing scoliosis-unique pain. Upon general research, that unique pain would include (but not be limited to) the following list:

1. Inguinal (groin/pelvic girdle; frequently related to the spinal Cobb Angle)
2. Cruralgia (symptoms radiating from the back typically to the front of the thigh)
3. Aching
4. Stiffness
5. Numb
6. Pain from back radiating to leg
7. Shooting

### SE-18 Possible Item Types

The pain scales reviewed predominantly utilized Likert style survey items. The frequent use of Likert style items might be limiting in their flexibility for individuals to describe the quality of their scoliosis pain in a meaningful way. At the same time, Likert items might be easier to deploy quantitatively especially with large sample sizes. Through other, more specific items types (like VAS or semantic differential style items) there could be an opportunity for a deeper investigation into the specific nature of adult scoliosis pain and then, as per the objective of this instrument, explore how the specific facets of that pain might or might not be impacted by exercise as a pain management strategy.

As revealed in a review of the literature, several researchers (Alanazi, et al., 2018; Zaina, et al., 202) noted a lack of quality data about the nature of scoliosis pain, making it challenging for researchers to understand if exercise is an appropriate pain management strategy recommendation.

So here we have established a cognitive dissonance of sorts. On one side, is the clinical viewpoint stating that chronic pain is mostly subjective. Although there is considerable hope that objective measures of brain function will soon replace subjective measures as the gold standard of pain measurement (Derbyshire, 2016). In light of this “gold standard,” it could be more useful, rather than attempting to understand each person’s individualized perception (quality) of their pain, to focus on frequency, intensity and how pain impacts QOL (as the sample scales have done).

But then what of the documented requests by researchers requesting more ample and higher quality data that specifically addresses the nature of scoliosis pain? Where is the middle ground here?

This is in part why the SE-18 aims to assess in adults with scoliosis the quality and nature of their pain while respecting its uniqueness. The ultimate goal is to gather enough such data to make a meaningful contribution to research that can make a determination as to whether exercise (including type, frequency, and amount) might be an effective pain management strategy for adults with scoliosis.

### Scale Differentiators

The goal is to create a scale that gathers data from a targeted population (see also “target sample”) of adults with scoliosis about their pain levels, exercise habits, and the effects of exercise on their pain levels and general functionality. This is especially important since many researchers reported a lack of thorough understanding of scoliosis pain and how it works, citing that lack of understanding as a barrier to recommending exercise as a potential clinically proven pain management intervention for scoliosis pain. To find out how scoliosis pain works and how it is impacted (or not) by physical activity, it would seem logical to create a survey instrument that measures that relationship.

The majority of research evidence reviewed addresses the relationship between exercise and curve reduction in adolescents with scoliosis but data appears to be lacking in the relationship between exercise and pain management in adults with scoliosis. Therefore, a goal of this instrument creation is to better understand the pain by learning more about long-term, day-to-day scoliosis pain through the eyes of a more targeted adult population, and how exercise impacts that pain.

## Methods

### Bracketing Subjectivity

As the sole project researcher, I feel it ethical to share how my subjectivity as an individual with chronic pain from scoliosis played a large role in the selection of this research topic. I have been living with chronic pain for over 35 years, due to extremely severe idiopathic scoliosis (initially diagnosed with a 116 degrees S-Curve) and subsequent Harrington Rod surgical correction (to 68 degrees) at age 19.

In 2017, having been unsuccessful in establishing a consistent movement habit and experiencing the effects of chronic pain on a daily basis, I essentially turned myself into an athlete (aerialist) as a way of committing to regular physical movement, conditioning, strength training, self-care and other tools of an athlete. Exercise, with time and consistency, ended up managing my pain level. Further, I am a former RN, BSN with a specialty area of orthopedics.

To manage my subjectivity to the best of my ability throughout this research project, I first conducted multiple, thorough literature reviews as a way of objectively obtaining as much knowledge and the most current research available on all aspects of my subject matter of interest. I also found that leaning into my personal knowledge of the lived experience of scoliosis, was helpful in the formulation of survey questions.

### Procedures

This project initially began in 2023 with a qualitative phenomenological investigation of generalized chronic pain and impact of exercise. The work from the start was grounded in multiple literature reviews on the subjects of chronic pain, scoliosis and scoliosis pain, and exercise in relation to both of those topics. The findings from these reviews as well as the creation of the construct, combined with subjective understanding of the research topic, provided the necessary guidance for item creation.

### Sample

The sample of survey respondents meeting the predefined criteria was formed from a combination of convenience, snowball, and volunteers sampling via direct email, social media, and in-person conversations/referrals. Surgical correction was not utilized as a factor in the sampling method. Individuals meeting the following criteria were recruited to take the survey via a public, online Qualtrics link:

1) Adults who have had scoliosis for 10 or more years.
2) Individuals experience or have experienced pain related to their scoliosis.
3) Individuals exercise, even occasionally.

Motivating the specific composition of the target survey respondents, was a desire to study pain, function, and quality of life in adult scoliosis patients. This is in opposition to the more frequent sample seen in this work, typically adolescent patients, mostly with smaller degrees of curvature (Cobb Angle) in relation to adults with scoliosis, and typically females with a mean age of 16 years old (Wong, et al., 2017).

As seen in *Table 1*, the resulting sample for this instrument (n=95 including records with missing data) was indeed composed of mostly female respondents, aged 20-30, 94% were diagnosed with scoliosis before age 21, the majority of Cobb angles (spinal curvatures) were self-reported to be 60 degrees or under, and most began experiencing pain 1 or more years post diagnosis.

**Table 1.**

|  |  |  |
| --- | --- | --- |
| Age Now | 21-30 y.o. 48%. | 51-60 y.o. 8% |
|  | 31-40 y.o. 23% | 61-70 y.o. 5% |
|  | 41-50 y.o. 15% | Over 70 1% |
| Gender | Female 90% | Male 10% |
| Age at Diagnosis | <12 y.o. 41% | 17-21 y.o. 5% |
|  | 12-16 y.o. 48% | >21 y.o. 6% |
| Degree of Curvature | 40% 10-40 degree curve |  |
|  | 45% 41-60 degree curve |  |
|  | 15% 61 degree or greater (severe scoliosis) |  |
| Onset of Pain | Before diagnosis, 17% | Since time of diagnosis, 11% |
|  | Within 1-5 years of diagnosis, 25% | Within 5-10 years of diagnosis, 21% |
|  | More than 10 years post diagnosis, 25% |  |

### The Survey

The SE-18 online survey was created in Qualtrics with a combination of multiple choice, Likert style items and “sliders” (Visual Analog Scale style). Face validity was tested by sending the initial survey draft to a panel of 3 experts in the subject matter of interest: 1) a physical therapist with her own practice who specializes in implementing scoliosis-specific Schroth exercises for pain and mobility management in adult scoliosis patients, 2) a primary care physician specializing in sports medicine and orthopedics, and 3) a highly respected orthopedic surgeon at Boston Children’s Hospital, Harvard Medical School Professor of Orthopedic Surgery, and respected scoliosis researcher.

Following the expert review of the first survey iteration for face validity, and implementation of expert feedback, the official pilot study was released to the public via the aforementioned sampling methods. The 18-question survey had 3 distinct areas of questioning: demographics/general information on the individual’s scoliosis condition, followed by in-depth questioning on the individual’s experience with scoliosis pain, mobility and other associated issues, and concluding with questions about if and how the individual currently exercises, if and how it impacts their scoliosis pain, and their attitude around the idea of exercise for scoliosis pain relief. The entire survey can be found for reference in Appendix A.

Following data collection via the survey instrument in Qualtrics, the raw data was downloaded in CSV as well as SPSS formats. All data analysis was done in IBM SPSS Statistics version 28. Analysis included descriptive statistics/frequencies, followed by reliability testing, and concluding with an exploratory factor analysis.

## Results/Analysis

### Raw Survey Data

The raw survey data picture of the data shows that most respondents experience some form of scoliosis and/or scoliosis-related pain daily. In items where participants were asked to rate their pain on a scale of 0-100, mean values showed that the highest number of respondents experience the most severe pain after standing/being on their feet for 1 hour or longer, with the second highest number of respondents experiencing their worst pain by the end of the typical day, rounded out by the percentage of respondents experiencing their highest pain levels after sitting or laying in one position for 1 hour or longer. Pain from holding the same body positions is a facet of a scoliosis also supported by the literature, which shows that those with back pain from scoliosis usually need to change position more frequently (Zaina, et al., 2023).

The highest pain location ratings were centered in the lower back, hips, and neck. In quality of pain ratings, the means trended closer together, with the highest values reflected in participants describing their pain as - dully/achy, tight, and sharp/stabbing. Respondents reported experiencing the most mobility and disability issues when not exercising regularly. Lying down/resting provided pain relief for the highest proportion of the sample, followed closely by stretching, and finally exercise.

Moving on to the exercise portion of the survey, 90% of respondents affirmed that they exercise, and those who exercise reported doing so more than once a week. Of those who exercise, 77% reported doing stretching, and 57% engage in some combination of strength training. 66% self-reported that exercise makes their scoliosis pain better, 18% report no change in their pain, and 16% reported that exercise makes their pain worse.

The majority of respondents stated that their pain restricts physical movement the most (mean of 42.62) when they are not exercising regularly. 79% were “extremely” open to the idea of exercise as a form of scoliosis pain relief. On a Likert style item where the responses were - yes, probably, maybe, and no - 69% believed that yes, they believed they would exercise regularly if exercise could help lessen their scoliosis pain. Of the types of exercises they would consider doing, 92% said they would engage in stretching, 84% would consider regular strength training, weights, and conditioning, and 76% said they would consider doing yoga for scoliosis pain relief.

The overall raw data picture reveals that most respondents experience pain, with over half reporting daily (chronic) pain, despite 40% of the sample having 10-40 degree curves. Recall how the literature found that pain occurs as a side effect of scoliosis when curvatures reach 30 degrees or higher (An, et al., 2023; Li, et al., 2021).

Also revealed in the raw data, is that most of the sample reported exercising regularly, and most would like to learn more about exercise as a form of scoliosis pain relief. These findings contrast with the multiple research findings that over half of scoliosis surgeons found no perceived value in prescribing exercise as a form of scoliosis pain relief.

### Descriptive Statistics

Frequencies were run on all survey questions, and a portion of those findings are seen here:

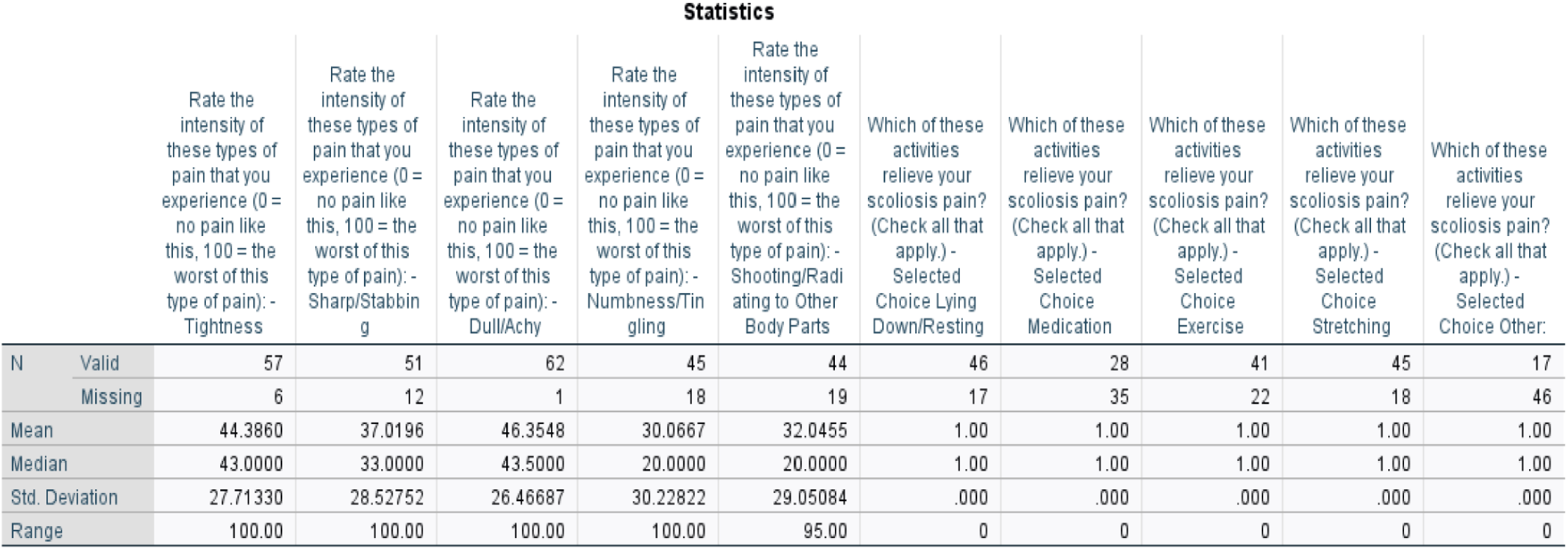

| Statistics |  |  |  |  |  |  |  |  |  |  |  |
| --- | --- | --- | --- | --- | --- | --- | --- | --- | --- | --- | --- |
|  |  | Rate the intensity of these types of pain that you experience (0 = no pain like this, 100 = the worst of this type of pain): - Tightness | Rate the intensity of these types of pain that you experience (0 = no pain like this, 100 = the worst of this type of pain): - Sharp/Stabbing | Rate the intensity of these types of pain that you experience (0 = no pain like this, 100 = the worst of this type of pain): - Dull/Achy | Rate the intensity of these types of pain that you experience (0 = no pain like this, 100 = the worst of this type of pain): - Numbness/Tingling | Rate the intensity of these types of pain that you experience (0 = no pain like this, 100 = the worst of this type of pain): - Shooting/Radiating to Other Body Parts | Which of these activities relieve your scoliosis pain? (Check all that apply.) - Selected Choice Lying Down/Resting | Which of these activities relieve your scoliosis pain? (Check all that apply.) - Selected Choice Medication | Which of these activities relieve your scoliosis pain? (Check all that apply.) - Selected Choice Exercise | Which of these activities relieve your scoliosis pain? (Check all that apply.) - Selected Choice Stretching | Which of these activities relieve your scoliosis pain? (Check all that apply.) - Selected Choice Other: |
| N | Valid | 57 | 51 | 62 | 45 | 44 | 46 | 28 | 41 | 45 | 17 |
|  | Missing | 6 | 12 | 1 | 18 | 19 | 17 | 35 | 22 | 18 | 46 |
| Mean |  | 44.3860 | 37.0196 | 46.3548 | 30.0667 | 32.0455 | 1.00 | 1.00 | 1.00 | 1.00 | 1.00 |
| Median |  | 43.0000 | 33.0000 | 43.5000 | 20.0000 | 20.0000 | 1.00 | 1.00 | 1.00 | 1.00 | 1.00 |
| Std. Deviation |  | 27.71330 | 28.52752 | 26.46687 | 30.22822 | 29.05084 | .000 | .000 | .000 | .000 | .000 |
| Range |  | 100.00 | 100.00 | 100.00 | 100.00 | 95.00 | 0 | 0 | 0 | 0 | 0 |

Due to the issue of missing data, a reduction in the number of valid cases was seen on most of the items found later in the survey, as compared to the initial Qualtrics reported sample size of 95. An informed decision was made, also in consultation with the project advisor, not to undergo time consuming missing data procedures for this initial pilot study. With additional time, following standard MCAR (Missing Completely at Random) practices, a data imputation procedure would have been performed. Entirely blank cases were removed, in addition to mostly incomplete cases where respondents completed only the initial 4 demographic/foundational information items of the survey. This resulted in a decrease from the initial 95 cases to 63.

### Scale Reliability

One purpose of statistical reliability analyses in survey instrument pilot studies like this, is to obtain feedback to improve the subsequent design of the instrument (Reh, et al., 2011). A survey pilot can help establish both reliability and validity, evaluate item consistency, identify possible biases, and assess the ability of the instrument to accurately collect the data it is designed to collect (Sparks Chart, 2024). In other words, it can help ensure that the survey measures what it promises to measure.

Reliability of the full 18-item scale was initially attempted in SPSS, however, due to the missing data issues, SPSS was unable to run a full scale analysis. After multiple trial and error attempts to run as much of the scale items in subsequently grouped runs as possible, reliability and item analyses on the following item combinations were run successfully: 6-9, 11-12, 14-15, 16-17.

For reliability coefficients, Cronbach’s standardized alphas were run to better account for this scale’s greatly disparate item variances. McDonald’s Omega was selected as a second, resilient reliability statistic, that can provide a more accurate reliability estimate when items are removed from a scale (as was the case here).

Finally, to maximize information gained in order to increase the quality of future iterations of this survey instrument, select Lambda reliability coefficients were obtained for each survey subscale depending on the nature of the data. For instance, L1 was run for each subscale, as a way of obtaining an intermediate coefficient. L4, Guttman’s split-half reliability, was included to see how effectively each subscale’s two halves measure the underlying construct. In the case of the subscale including items 6-9, L6 was interpreted because, as indicated for this coefficient, upon examination, inter-item correlations were low compared to squared multiple correlations.

**Table 2.**

| Item Range | Distinct Construct | Valid/<br>Excluded Cases | Cronbach's<br>(Standardized)<br>Alpha | McDonald's<br>Omega | Lambda | Interpretation |
| --- | --- | --- | --- | --- | --- | --- |
| 6-9 | Frequency,<br>intensity,<br>quality of<br>pain | 21/42 | .927 | .948 | 1: .893 | A reliable subset of items for the most part, with items that would appear to measure what the distinct construct claims they measure. But this is also a smaller subset of data in number of cases.<br>However, since Q7-9 were VAS/slider items generating many data points, there was a lot to work with here in generating a strong reliability analysis with high coefficient values. |
|  |  |  |  |  | 4: .876<br>(Guttman) |  |
|  |  |  |  |  | 6: .996 |  |
| 11-12 | Assessing<br>exercise<br>habits | 61/2 | .588 | n/a (fewer<br>than 3<br>variables) | 1: .164 | A large percentage of valid cases, but the reliability statistics indicate that these 2 items might not be measuring the same construct. "Do you exercise (yes/no)?" and "How frequently (m. choice)?" are admittedly nonspecific queries. |
|  |  |  |  |  | 4: .327 |  |
|  |  |  |  |  | 6: .309 |  |
| 14-15 | Exercise and<br>scoliosis<br>pain/disability | 50/13 | .756 | .853<br>(More than 3<br>variables as<br>Q15 was 3<br>sliders-in-one<br>item.) | 1: .542 | Another 2-item construct but unlike the pairing above, one of the items here was a VAS/slider, actually 3 sliders in 1 (Q15), thereby providing more data for reliability analysis than the 11-12 subscale items above.<br>"Does exercise make your scoliosis pain... (likert style - better, same, worse)" & the 3-in-1 VAS addressing |
|  |  |  |  |  | 2: .804 |  |
|  |  |  |  |  | 4: .856 |  |
|  |  |  |  |  |  | how much pain restricts mobility, showed a higher level of subscale reliability. |
| 16-17 | Futurecasting<br>- assessing<br>willingness to<br>exercise. | 62/1 | .782 | n/a (fewer<br>than 3<br>variables) | 1: .362 | The final subset and once again a 2-item one, had the highest number of valid cases yet. While McDonald's Omega was not able to be calculated, Cronbach's Standardized Alpha and Guttman's Split Half Reliability indicated that these 2 items are likely measuring the same or a similar latent construct.<br>Q16 asks how open the person is to exercise as scoliosis pain relief (Likert style) and Q17 queries if the person believes regular exercise could be a form of scoliosis pain relief (also Likert style responses). |
|  |  |  |  |  | 4: .724 |  |

### Item Analysis

Item-Total Statistics, specifically Correlated Item-Total Correlation and Cronbach’s Alpha if Item Deleted, were run for each subset of the scale wherein the following observations were made.

### Items 6-9: Scoliosis Pain & Disability

The removal of most individual items, made the scale subset slightly more reliable (initial Cronbach’s Standardized Alpha, .927). One interpretation of this finding could be that each of these items has a similar amount of strengths and weaknesses contributing to the full survey instrument. This might also indicate that, since normally a shorter scale would decrease, not increase reliability, one or more of these items would be introducing noise or redundancy to this items subset.

**Item-Total Statistics**
|  | Scale Mean if Item Deleted | Scale Variance if Item Deleted | Corrected Item-Total Correlation | Squared Multiple Correlation | Cronbach's Alpha if Item Deleted |
| --- | --- | --- | --- | --- | --- |
| How frequently do you experience scoliosis pain? | 732.2381 | 161407.290 | -.573 | .963 | .946 |
| Rate your overall scoliosis pain on a typical day (0 = no pain, 100 = worst pain):<br>- When you wake up in the morning: | 698.8571 | 143265.129 | .731 | .996 | .939 |
| Rate your overall scoliosis pain on a typical day (0 = no pain, 100 = worst pain):<br>- By midday: | 697.3810 | 147634.448 | .713 | .922 | .939 |
| Rate your overall scoliosis pain on a typical day (0 = no pain, 100 = worst pain):<br>- By the end of the day: | 676.7619 | 143542.290 | .779 | .994 | .938 |
| Rate your overall scoliosis pain on a typical day (0 = no pain, 100 = worst pain):<br>- After standing for an hour or more: | 671.5714 | 147225.357 | .566 | .936 | .941 |
| Rate your overall scoliosis pain on a typical day (0 = no pain, 100 = worst pain):<br>- After sitting or laying in one position for an hour or more: | 683.9524 | 143328.148 | .694 | .979 | .939 |
| Rate your scoliosis pain in the following areas on a typical day (0 = no pain, 100 = worst pain): - Upper Back | 696.9524 | 141009.448 | .772 | .997 | .938 |
| Rate your scoliosis pain in the following areas on a typical day (0 = no pain, 100 = worst pain): - Middle Back | 697.0952 | 141028.790 | .837 | .969 | .937 |

In reviewing item correlations, item Q6, “How frequently do you experience scoliosis pain? (Likert style responses)” is negatively correlated with the other items, 7-9. This makes intuitive sense, as items 7-9 are all VAS/slider items, allowing respondents to pinpoint various aspects of their pain, on a 0-100 scale. It may be that item 6 is too broad and disconnected from the more specific, ratings-enabled conversation about scoliosis pain in items 7-9.

Another lower item correlation was one of the sliders from Q9, asking respondents to evaluate the quality of their scoliosis pain, with various adjectives offered, and again on a scale of 0-100. The slider asking respondents to rate their “dull/achy” pain correlated lower than the rest of this subset, at .489. This might indicate that the categorization of scoliosis pain as “dully/achy” might be less inherently similar to various other characterizations of scoliosis pain reflected in items 6-9.

Overall for this subset (within a subset) of items 7-9, the majority of items correlated to the subset at .7 or higher, a possible statistical indicator that items 7-9 together measure more of the same latent variable than items 6-9 measure together.

### Items 11-12, 14-15, 16-17: Scoliosis Pain & Exercise

There were far more item and scale reliability issues with the exercise-focused subsets of the survey instrument. Based on observations thus far about the item types yielding the highest reliability (VAS/sliders), these issues could be related to the inclusion of only one such item, Q15, allowing respondents to rate how pain restricts their physical movement - on a typical day and when exercising vs. not exercising regularly. Following are statistical assessments from the item reliability analysis of the exercise-related items.

### Inter-Item Correlation Matrix

● At .802, the amount of pain restricting mobility when the individual is exercising regularly, was highly correlated with the extent to which pain interferes with mobility on an average day (0-100 slider VAS items). In other words, regular exercise meant less pain and better mobility.
● A negative correlation of -.069 was found in the 2-item scale subset - “Does exercise make your scoliosis pain better/same/worse?” and a 0-100 slider item assessing the relationship between pain and mobility on a typical day, suggesting that this specific pair of items might not be reflective of the same latent construct.
● Finally, the pair of questions asking participants if they believed they would exercise regularly if it helped relieve their scoliosis pain, and how open they are to the idea of exercise as scoliosis pain relief, correlated with one another at .642, suggesting a possible latent construct match.

### Item-Total Statistics

● Looking at item-total correlations, with a fairly low correlation of .312, “Does exercise make your scoliosis pain better/same/worse?” (items 14-15 subset) appears to be less related to measuring how pain impacts mobility in relation to exercise. Cronbach’s Alpha increases to .809 (from .756) when the latter item is removed from this admittedly micro subset. Similar to prior observations on other items experiencing this statistical effect, this could indicate noise or redundancy on the part of this item.
● Alternately, Q15 (a slider incorporating 3 sub questions), addressing pain and mobility on a typical day vs. exercising regularly vs. not doing so, shows a high item-total correlation at .835. Cronbach’s Alpha dips from .756 to .417 when this item is removed, revealing another strong VAS/slider type item likely with interconnected ideas being measured.
● The Q15 slider questions matching up pain, mobility, and exercise showed a high item-total correlation of .835 for “exercising regularly” and .660 for “not exercising regularly.” It makes theoretical sense that the affirmative and negative of the same question would correlate highly. Cronbach’s Alpha goes from .756 to .679 “when exercising regularly” is removed, and from .756 to .565.

### Reliability Analysis Summary

#### Item Type Matters

Based on initial assessment of these scale reliability results, VAS/slider items seemed to perform much better than Likert style, multiple choice, and dichotomous style items (in that descending order). It could be that the specificity offered in rating various aspects of scoliosis pain, mobility, and exercise experienced by respondents, was better accomplished using VAS/slider items.

#### Novice Item Writing

The scale and item reliability analyses also reflect item writing room for improvement. Even prior to the Exploratory Factor Analysis, many of the reliability results showed that the majority of the “non-VAS/slider” items measured in subsets, were not particularly well-related ideas, possibly not measuring the same latent variable(s).

#### General vs. Specific Questions

Lack of specificity in item writing might have also played a role in scale subset reliability issues. More generally worded items, particularly either dichotomous or brief multiple choice ones, might not have been as effective in uncovering latent constructs and in creating a reliable scale. This theory is also supported by a large amount of additional, qualitative-style data generated by the instrument, through the “other” option on many items. This data could indicate a participant preference for describing their experience with scoliosis and exercise in more specific detail.

### “Other”

As noted in the previous section, a great deal of information expanding and elaborating on the quantitative findings of this pilot study survey was provided via a small number of open-ended/”other” item responses.

### Q10: Which of these activities relieve your scoliosis pain, check all that apply - “Other” (28%)

**Q10: Which of these activities relieve your scoliosis pain, check all that apply - “Other” (28%):**
|  |
| --- |
| Other: |
| Heating pad |
| Walking |
| Massage; heat (heating pad, hot tub) |
| Hot shower am daily |
| Massage |
| Heating pad and ice |
| Postural reeducation physiotherapy |
| yoga |
| Marajuana and Tylenol/Aleve/Advil, Lidocaine Patches, Tens Unit, Physical Therapy, Nerve Blockers, Massages |
| Not really an 'other', but I thought it would be important to note that while all the checked boxes do provide some relief, it's temporary and sometimes worsens the pain as well. |
| Hear |
| Relief in the short term is hard massage or vibration on the bones of my lower spine |
| Changing positions |
| Back rub/massage |
| Heating pad |
| Massage |

**Q13: What kinds of exercises do you do, check all that apply - "Other" (41%):**

### Q13: What kinds of exercises do you do, check all that apply - “Other” (41%)

|  |
| --- |
| Other |
| Swimming |
| Walking |
| Walking, bicycling, aqua fitness |
| Playing with my kids - tennis currently |
| Walking |
| Rowing |
| Walk |
| Walking, gardening |
| swimming, walking |
| I used to be really athletic and always worked out in HS and the pain wasn't as bad as now that I've quit working out |
| Specific physiotherapy for scoliosis (Italian SEAS); proprioception exercises; pilates with focus on mobility exercises |
| Walking |
| General Everyday Actions, walking, leaning, moving a lot. |
| Daily walks around 4-5km |
| Schroth method |
| Aerial silks/lyra |
| Climbing |
| Physiotherapy, I'm not sure whether that falls under pilates/yoga, but it's basically stretching and core exercises and exercises to correct posture. |
| Tennis Walking |
| Physical therapy, walking |
| swimming |
| Spin (cycle) classes, walking, swimming, |
| spin class, running |
| Walking 4-5 miles daily |
| Biking, walking, stair stepper |

### Q18: Which exercises would you consider doing if it would relieve your pain in the long-term, check all that apply - “Other” (15%)

**check all that apply - "Other" (15%):**
|  |
| --- |
| Other |
| Swimming |
| hiking, walking outside |
| Walking |
| Walking for an hour |
| Walking |
| The ones mentioned before |
| Walking |
| Cannot run due to unrelated hip deformity. |
| Climbing |

Did adults with scoliosis who participated in this pilot study see the opportunity for open ended “other” questions as an opportunity to share their unique experiences with scoliosis?

The forthcoming exploratory factor analysis will attempt to uncover the latent factors being measured by the scale subsets. Ideally, all data revealed by this pilot study, open and close ended alike, will contribute to the further development of this survey instrument.

### Exploratory Factor Analysis

Exploratory Factor Analysis (EFA) explores the structure of observed data to determine whether select latent variables or constructs are responsible for multiple individual variables, in this case, how individual survey items reflect the construct of a larger psychometric instrument (Schreiber, 2021; Columbia Mailman School of Health, 2024). EFA is important in the pilot testing of survey instruments like this one, as a manner of examining theory, testing scale validity, and reducing scale structure for future analysis (Scispace, 2024).

An EFA of the full 18-item scale was initially attempted in SPSS, however, due to similar missing data issues as occurred in the reliability analysis, SPSS was unable to run a full scale analysis. An intentional choice was made to break the scale into the same subsets (and in the case of the “rate the…”/VAS/Slider questions, subsets within subsets) as were run in the reliability analysis: items 6-9, 11-12, 14-15, and 16-17.

A variety of factor extraction and rotation combinations were run on each of the scale subsets, but the clearest, most consistent factor solution, that was best supported by theory and research, was found in the Principal Axis Factoring (PAF)/Varimax combination.

PAF is a frequently used extraction method with no distributional assumptions, robust in unequal factor loadings and smaller sample sizes, both beneficial in a survey pilot study like this (Grieder, 2022). For factor rotation, Varimax, an orthogonal rotation, is also a common technique, which makes high loading factors load higher, and lower loading factors load lower, for more definitive results analysis (Akhtar-Danesh, 2023).

### EFA on Items 6-9: Nature of Scoliosis Pain

Communalities were first explored on survey items 6-9, the most robust subset of the scale, which explored the respondents’ description of the nature and impact of scoliosis pain. The top communalities, defined as those ranking .8 or higher, in order from the most variation accounted for by latent traits, to the least explained were as follows:

1. Rate intensity of certain type of pain - numbness/tingling: .902
2. Rate your scoliosis pain in this area on typical day 0-100 - hips: .895
3. Rate your scoliosis pain in this area on typical day 0-100 - legs: .863
4. Rate your overall scoliosis pain on a typical day - first thing a.m.: .858
5. Rate your scoliosis pain in this area on typical day 0-100 - upper back: .850
6. Rate your overall scoliosis pain on a typical day - end of day: .808
7. Rate your scoliosis pain in this area on typical day 0-100 - middle back: .806
8. Rate your overall scoliosis pain on a typical day - same position: .804

On the other end of the spectrum, the two lowest communalities were: “How frequently do you experience scoliosis pain (multiple choice)?” at .461 and “rate your scoliosis pain by location - other” at .452.

Next, factor extraction and rotation were performed to assess the number of potential factors that might explain survey items 6-9.

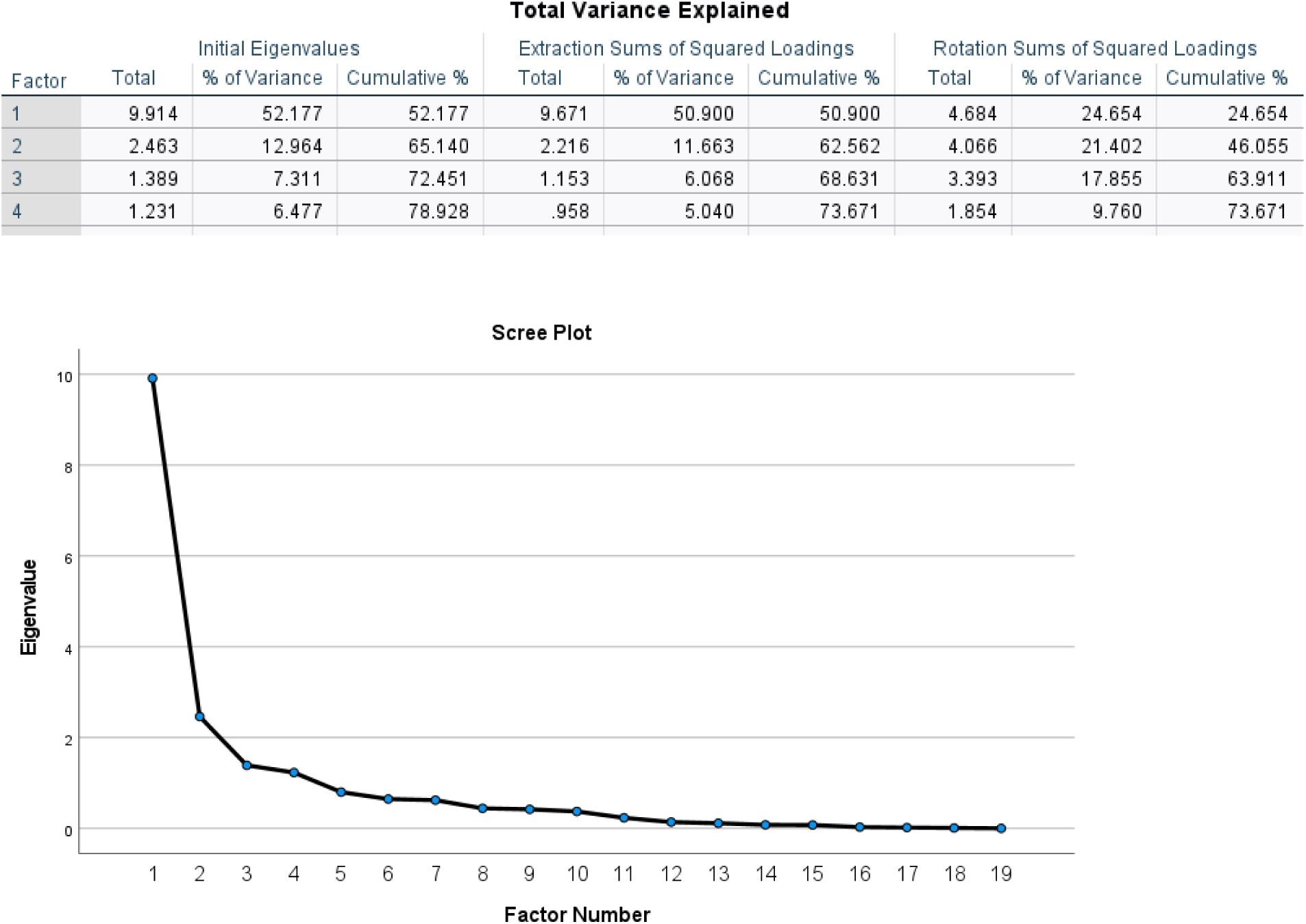

Based on the Eigenvalue cutoff value of 1.00, the primary method of extraction/rotation, PAF/Varimax found a 4-factor solution (73.6% of variance explained). However, upon review of the scree plot, the extracted factors, noting that the majority of variables loaded onto 3 variables at .3 or higher (63.9% of variance explained), and leading with theory/research, a 3-factor solution, as seen in the following table, made the most sense.

| 1 | 2 | 3 |
| --- | --- | --- |
| Rate your overall scoliosis pain on a typical day - first thing a.m | Rate intensity of certain type of pain - radiating to other body parts | Rate your overall scoliosis pain on a typical day - by end of day |
| Rate your overall scoliosis pain on a typical day - midday (cross loaded with #3) | Rate your scoliosis pain in this area on typical day 0-100 - arms | Rate your scoliosis pain in this area on typical day 0-100 - upper back (cross loaded with #2) |
| Rate your overall scoliosis pain on a typical day - standing over an hour | Rate your scoliosis pain in this area on typical day 0-100 - legs (cross loaded with #1) | Rate your scoliosis pain in this area on typical day 0-100 - middle back |
| Rate your overall scoliosis pain on a typical day - same position (laying or sitting) | Rate intensity of certain type of pain - numbness/tingling | Rate your scoliosis pain in this area on typical day 0-100 - neck (cross loaded with #2) |
| Rate your scoliosis pain in this area on typical day 0-100 - lower back | Rate intensity of certain type of pain - sharp/stabbing | Rate your scoliosis pain in this area on typical day 0-100 - other (LOW loading) |
| Rate your scoliosis pain in this area on typical day 0-100 - hips |  | Rate intensity of certain type of pain - tightness (cross loaded with #1) |
| *Themes: Morning stiffness, holding the same body positions, lower back and hip pain, all correlated with mobility issues.* | *Themes: Pain, numbness, tingling, sharp, stabbing, radiating to limbs. Potential neurological themes.* | *Themes: End of day, fatigue, upper back, neck, tightness from overuse. Symptoms that speak to stress and body tension.* |

In additional extraction/rotation combinations, PAF/Promax showed a pattern matrix that was an almost perfect match to PAF/Varimax, which via the Eigenvalue cutoff was consistent with a 4 factor solution. It was also consistent with the PAF/Varimax scree plot and theory interpretation of 3 factors. In an Alpha (assumes that all variance among variables is shared)/Promax combination, nearly all variables loaded onto factor 1.

Returning to the primary PAF/Varimax combination, the only survey item that failed to load was “How frequently do you experience scoliosis pain (multiple choice)?” also as noted in prior results, a previously problematic item. Once again, the theory on such nonspecific items, is that they do not provide enough information to be definitively reflective of a latent trait.

### EFA on Items 11-12: Assessing Exercise Habits

The extractions on the exercise-related 2-survey item subscales were more straightforward, beginning with the item 11-12 combination: Do you exercise/If you do, how frequently? Once again using a PAF/Varimax combination, both variables showed communalities of .415, indicating an equal amount of variation accounted for by latent traits, both with the same level of extraneous variables. A ULS (used when the researcher does not want to make distribution assumptions but statistical efficiency can be sacrificed)/Promax showed .173 communalities, and Alpha/Promax at .415.

Looking at the suggested number of factor solutions based on an Eigenvalue cutoff of 1.00, all 3 extraction/rotation combinations found a 1-factor solution (PAF/Varimax with 42% variance explained, ULS/Promax at 71%, Alpha/Promax at 42%). Both variables extracted at .644 in an unrotated matrix, as a rotated factor matrix could not be produced due to too few variables. These results would seem to indicate that the 1 latent trait behind this 2 item scale subset, might be exercise.

### EFA on Items 14-15: Exercise, Scoliosis Pain and Mobility

This subset pairing provided slightly more information, due to Q15 being a 3-in-1 rating/VAS/slider question (measuring how much pain restricts mobility in a typical day, when not exercising, and when exercising). Q14 asked if exercise makes respondents’ scoliosis pain better, the same, or worse. When these 2 items and their sub items were combined, 4 variables were extracted and communalities were as follows:

1. Q15: Pain/Mobility/Exercising Regularly: .904
2. Q15: Pain/Mobility/Average Day: .875
3. Q14: “Does exercise make your scoliosis pain - better, the same, worse”: .597
4. Q15: Pain/Mobility/NOT exercising regularly”: .592

Based on a review of eigenvalues (1.00 or higher) and scree plots, using the PAF/Varimax, PAF/Promax, Alpha/Promax, and ULS/Varimax, once again leading with PAF/Varimax, it was found that a 2-factor solution explained 74% of variance.

| Factor | Initial Eigenvalues |  |  | Extraction Sums of Squared Loadings |  |  | Rotation Sums of Squared Loadings |  |  |
| --- | --- | --- | --- | --- | --- | --- | --- | --- | --- |
|  | Total | % of Variance | Cumulative % | Total | % of Variance | Cumulative % | Total | % of Variance | Cumulative % |
| 1 | 2.407 | 60.185 | 60.185 | 2.219 | 55.479 | 55.479 | 1.542 | 38.558 | 38.558 |
| 2 | 1.129 | 28.236 | 88.420 | .748 | 18.711 | 74.190 | 1.425 | 35.632 | 74.190 |
| 3 | .326 | 8.159 | 96.579 |  |  |  |  |  |  |
| 4 | .137 | 3.421 | 100.000 |  |  |  |  |  |  |

Extraction Method: Principal Axis Factoring.

Factors extracted as follows:

Extraction Method: Principal Axis Factoring. Factors extracted as follows:
| 1 | 2 |
| --- | --- |
| Pain/mobility/average day | Does exercise make your scoliosis pain - better, same worse (~66% said better) |
| Pain/Mobility/NOT exercising | Pain/Mobility/Exercising Regularly |
| *Themes: State of scoliosis pain when not exercising/sedentary* | *Themes: State of scoliosis pain when exercising.* |

### EFA on Items 16-17: Willingness to Exercise

Finally, EFAs were run on survey items 16 & 17, both multiple choice items addressing how open respondents were to the idea of exercise as scoliosis pain relief, and if they believed regular exercise would help them relieve scoliosis pain.

Both variables showed .641 communalities, leading once again with PAF/Varimax, and Eigenvalues confirmed a 1-factor solution similar to the earlier item pairing, with that factor most likely being exercise.

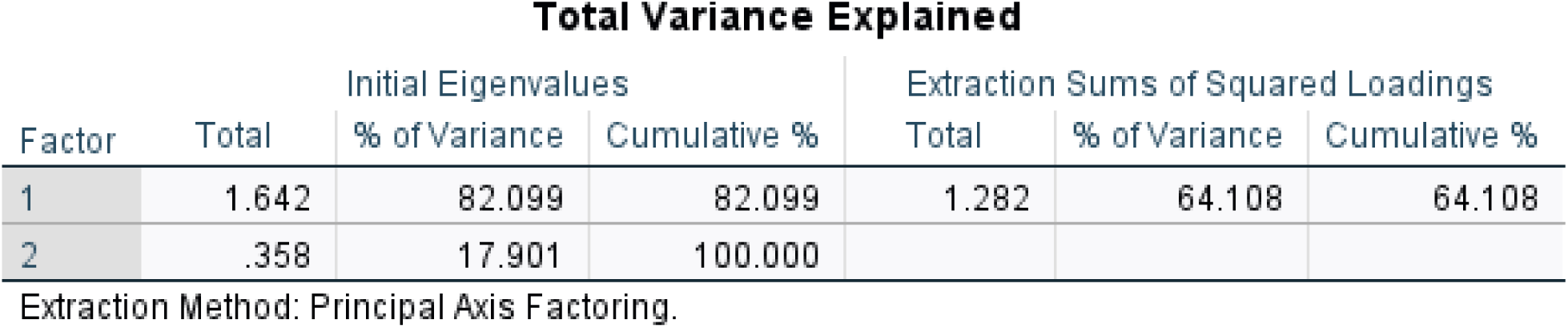

## Discussion

### Item Writing: Learning Curves Ahead

Item writing, when done well, can be a long, involved, and iterative process. The correlation between well-written, specifically focused questions, and scale reliability and factor construct clarity, became clear throughout the data analysis phase of this study.

Another “meta” lesson learned was the importance of matching the best item type to the question being asked. It was apparent during data analysis, that the item type which generated the the most robust raw data, the most reliable scale subset coefficients, and the most information about the possible latent construct behind the survey, was the “rate your pain/mobility/etc… on a scale of 0-100” items. These “VAS/Slider” items admittedly generated more data, as each one contained multiple questions in one.

VAS/Slider items showed highly correlated data which, in most cases, reflected an underlying latent construct. What might account for the lack of ambiguity in this data describing the scoliosis pain experience, is the motivation of the people who provided the data. It is possible that respondents might have welcomed the opportunity to respond to a quantitative item, but in a nuanced way where they could describe facets of their unique scoliosis pain. As it turned out, the greater the opportunity to describe the facets of their scoliosis pain experience, the better that lived experience is described.

The differences in statistical results between VAS/Slider items and the other item types, might be the difference between narrow questions that participants had been asked before about their scoliosis experience, and the opportunity to delve in deeper, sharing more detail, but still in a quantitative manner.

Also, as discussed in the results section, the “other” open-ended options on some questions, while not great for reliability or EFA purposes, yielded a generous batch of rich data from which to craft future iterations of the instrument. Overall, the amount and quality of raw data, along with the results of the statistical analyses, have laid a promising foundation for future work on the SE-18.

### Exercise: Left To Their Own Pain Coping Devices

The reliability and EFA results on the exercise item subsets were less impressive than the pain-related subsets. But the exercise items as a whole did yield some interesting theoretical seeds, ripe for further exploration, about the target population of adults with scoliosis who experience pain and take part in exercise, even occasionally. One of those seeds, obtained through the “other” open-ended response questions is that, when faced with great, enduring, long-term struggle, like chronic pain - people will find their own solution.

The qualitative data obtained from the open-ended “other” items revealed that many of the survey respondents have tried or are trying a spectrum of possible solutions in order to manage their pain. This was especially evident in the laundry list of types of exercise written in by respondents. Far beyond “traditional exercise methods,” a handful of respondents listed scoliosis specific exercise as part of their pain control regimens, something also supported by the literature. This reinforces the innate value of a survey pilot instrument, providing quantitative statistical analysis on the reliability and latent construct powering the scale, as well as meta analysis of the survey construction. When open-ended questions are included, researchers can gain even more detailed information to craft future iterations of the instrument.

### Latent Traits Worthy of Further Exploration: More Than Back Pain

One of the most compelling statistical results of this pilot study was the potential 3 factor solution emerging from the first, pain-focused scale subset, items 6-9 (recall items 7-9 were VAS/slider items generating an abundance of data). The proposed 3 factor solution is in regard to the nature of scoliosis pain: Mobility Issues, Neurological Issues, and Body Tension/Stress. What follows is reasoning for each possible latent trait based on the EFA, literature review, and theory.

1) Mobility Issues: variables extracted indicated possible morning mobility issues, discomfort from holding the same body postures, and lower back and hip pain correlated with mobility issues. The research confirms a connection between scoliosis and hip issues. Roberts (2023) found that the biomechanical impact on the hips and pelvis due to the asymmetry that is characteristic of scoliosis, especially over time, can cause degenerative changes and stress to the hip joints.
2) Neurological Issues: variables extracted described the quality of scoliosis pain, with descriptors such as numbness, tingling, sharp, stabbing, and pain radiating to the limbs, all of which could describe neurological symptoms. Smith et al. (2008) found that neurological symptoms, not unlike the ones featured on this latent factor, are common in adults with scoliosis.
3) Body Tension/Stress: variables loading on factor 3 could reflect body fatigue and tension at the end of one’s day, particularly with variables describing pain in the upper back and neck, as well as describing their scoliosis pain as “tightness,” possible signs of stress and body tension.

As for the EFA on the exercise related scale subsets, the factor verdict on most of the 2 survey question subsets seemed to be one single factor - exercise. For the item 14-15 pairing, the two factors also appeared straightforward - 1) state of scoliosis pain when not exercising, 2) state of scoliosis pain when exercising. Together, the EFAs on the exercise items, suggest that more extensive item writing work needs to be done on these items in particular, in future iterations of this instrument, to better identify the latent construct behind the scoliosis pain and exercise relationship.

### Research Limitations

One limitation of this pilot study was the convenience sample of respondents recruited through the researcher’s contacts along with outreach to strangers via social media and other online means, using a publicly issued survey URL. This also meant that prescreening of participants was entirely self-reported.

Second, a need for more time for higher quality, iterative item writing was thought to be a limitation in this pilot study, in that it prevented full scale analyses, which might have yielded more robust, consistent raw data.

The third limitation was the large amount of missing data that likely affected the quality of analysis. Finally, the lack of a post-survey focus group to obtain feedback on the survey itself, was a limitation in terms of identifying improvements for future survey iterations.

## Conclusion

### Project Goal

The goal of this project was to develop a survey instrument, to the pilot study phase, that gathers subjective data about the impact of exercise/movement on pain levels and functionality in adults with chronic pain from scoliosis.

Key findings of this pilot study included:

● High reliability coefficients in the portion of the instrument which measured the nature of scoliosis pain in adults with scoliosis who experience or who have experienced pain, and who exercise at least occasionally.
● The exercise-themed items of the instrument showed lower reliability, and may require item writing improvement in order to obtain more descriptive and meaningful data in future survey iterations.
● A 3-factor solution on the nature of scoliosis pain in respondents was identified using a variety of extraction and rotation techniques. Those 3 factors were: Mobility Issues, Neurological Effects, and Body Stress/Tension. For exercise, a tentative 2-factor solution making the distinction between “pain experience while exercising regularly” and “pain experience while not exercising regularly” was identified.
● An overall pattern in the data, including qualitative questions inviting respondents to elaborate on their survey responses, indicated that respondents embraced opportunities to describe their unique experience with scoliosis pain and exercise vs. close-ended item types (multiple choice, dichotomous, Likert).

### Implications

To achieve better results, the survey instrument will need to be further developed and refined based on the findings of this pilot study. Closer attention should be paid to creating a scale wherein the items are measuring the same construct, noise and redundancy is reduced within items, a new group of respondents is more carefully curated with pre screening techniques, and survey “meta” follow-up occurs for future survey refinement.

Overall however, the survey instrument performed better and yielded a greater amount of useful information than was anticipated. A detailed understanding was gained of how respondents experience scoliosis pain and associated physical issues, a possible latent construct summarizing their experience, and how they view exercise as a potential method of managing their pain.

### Research Significance

Research like this, that demonstrates the potential value of scoliosis-specific exercise strategies, could be applied as long-term pain management in adults with scoliosis with static spinal curvatures. Further, if shown to be effective, exercise strategies could be recommended to newly diagnosed scoliosis patients as long-term, proactive pain management solutions.

Scoliosis constants like pain management, quality of life, and improved functionality, should be theoretically separated from initial, short-term clinical goals like curve improvement. This approach, of separating pain and mobility symptoms from degree of curvature, presents an opportunity to focus on a population of scoliosis patients who experience compounding pain and decreased quality of life over the long term.

The longer term goal of this instrument, beyond this pilot study, is to study the relationship between scoliosis pain and exercise in the sub-population of individuals with scoliosis who ostensibly have the greatest need for pain management strategies to improve their quality of life.

### Final Takeaways

The results of this pilot study suggest that the experience of scoliosis pain is more complex than “back pain,” and exercise as a potential form of pain management is more complicated than “go exercise and you’ll feel better.” Like other types of chronic pain, adults with scoliosis are eager to share the details of *their* unique experience, and when it comes to finding relief, they are willing to forge their own path until a better one is provided for them. The purpose of this work has always been and remains, to help these individuals forge a research supported path or paths that can be made available to all who live with adult scoliosis pain.

### Future Research Directions

● **SE-18 Rev 2.0:** The results of this project, as well as a revisiting of initial expert validity feedback and a continuous review of the literature, will be utilized to create a revised version of the instrument for a new research study..
● **Multiple Survey Versions:** Also being considered is breaking out this survey into 2 different surveys, one targeting scoliosis patients who have undergone corrective surgery, and the other survey targeting those who have not had corrective surgery.
● **Improved Content Validity:** A new and more extensive round of expert validity feedback will be solicited for each of those two surveys, from more experts, and using a more thorough process. For example, experts will be asked to rank in order, which items are most reflective of the construct down to least reflective.
● **More Rigorous Sampling:** A more comprehensive and longer sampling and survey distribution period will also be instituted, with better pre-screening (for higher validity) of respondents, and improved tracking and communication procedures.
● **Qualitative Meta Data:** A Zoom focus group of randomly selected participants will be utilized to obtain feedback on the survey experience and items.
● **Missing Data Procedures:** Once data from those surveys is collected (using a higher tier of Qualtrics plan for better data collection and reporting features), missing data will be imputed using a MCAR or other appropriate missing data procedure, for higher quality data to be analyzed.
● **Improved Statistical Analyses:** Reliability and EFA analyses like the ones run in this pilot study will once again be utilized for data analysis. Ideally, with improved item writing, the entire scale will be successfully analyzed at once, rather than analyzing subsets of items.
● **High Quantity and Quality of Research on This Topic:** Also of interest for a longer-term investigation, a study of longitudinal vs. short-term exercise programs, either as a randomized control trial or quasi-experimental research design. There might be some potential with single-case and multiple baseline designs as well, applying and removing different types of exercises as varying treatment conditions. Overall, a higher quantity of more rigorous designs are needed to create more robust, generalizable evidence for clinicians in the field.

## Data Availability

All data produced in the present study are available upon reasonable request to the authors.

## References

Ahonen, M., Syvänen, J., Helenius, L., Mattila, M., Perokorpi, T., Diarbakerli, E., Gerdhem, P., & Helenius, I. (2023). Back pain and quality of life 10 years after segmental pedicle screw instrumentation for adolescent idiopathic scoliosis. Spine, 48(*10*), 665–671. 10.1097/brs.0000000000004641

Akhtar-Danesh N. (2023). Impact of factor rotation on Q-methodology analysis. PLoS One. Sep 1;18(9):e0290728. doi: 10.1371/journal.pone.0290728.

Alanazi, M. H., Parent, E. C., & Dennett, E. (2018). Effect of stabilization exercise on back pain, disability and quality of life in adults with scoliosis: a systematic review. European Journal of Physical and Rehabilitation Medicine, 54(5). 10.23736/s1973-9087.17.05062-6

An, J. K., Berman, D., & Schulz, J. (2023). Back pain in adolescent idiopathic scoliosis: A comprehensive review. Journal of Children’s Orthopaedics, 17(*2*), 126–140. 10.1177/18632521221149058

Araújo, M. E. A., Silva, E. B., Mello, D. B., Cader, S. A., Salgado, A. S. I., & Dantas, E. H. M. (2012). The effectiveness of the Pilates method: Reducing the degree of non-structural scoliosis, and improving flexibility and pain in female college students. Journal of Bodywork and Movement Therapies, 16(2), 191–198. 10.1016/j.jbmt.2011.04.002

Asher, M., Min Lai, S., Burton, D. & Manna, B. (2003). The reliability and concurrent validity of the scoliosis research society-22 patient questionnaire for idiopathic scoliosis. Spine 28(*1*), 63–69.

Bess, S., Boachie-Adjei, O., Burton, D., Cunningham, M., Shaffrey, C., Shelokov, A., Hostin, R., Schwab, F., Wood, K., Akbarnia, B. & the International Spine Study Group (2009). Pain and disability determine treatment modality for older patients with adult scoliosis, while deformity guides treatment for younger patients. Spine 34(*20*), 2186–2190. 10.1097/BRS.0b013e3181b05146

Blum, C. L. (2002). Chiropractic and pilates therapy for the treatment of adult scoliosis. Journal of Manipulative and Physiological Therapeutics, 25(4), E1–E8. 10.1067/mmt.2002.123336

Chen, C., Xu, J., & Li, H. (2024). Effects of Schroth 3D exercise on adolescent idiopathic scoliosis: a systematic review and meta-analysis. Children, 11, 806. 10.3390/children11070806

Chen, Y., Zhang, Z., & Zhu, Q. (2023). The effect of an exercise intervention on adolescent idiopathic scoliosis: a network meta-analysis. Journal of Orthopaedic Surgery and Research, 18:655. 10.1186/s13018-023-04137-1

Columbia Mailman School of Health. (2024). Exploratory factor analysis. https://www.publichealth.columbia.edu/research/population-health-methods/exploratory-factor-analysis

Dahlbäck, A., Andréll, P. & Varkey, E. (2023). Reliability and aspects of validity of the Swedish version of self-efficacy for exercise scale for patients with chronic pain, Physiotherapy Theory and Practice, 39:*1*, 163–173, 10.1080/09593985.2021.1999356

Derbyshire, S.W.G. (2016). Pain and the dangers of objectivity. In: van Rysewyk, S. (eds) Meanings of Pain. Springer, Cham. 10.1007/978-3-319-49022-9_2

Djurasovic, M., Glassman, S., Sucato, D., Lenke, L., Crawford, C., & Carreon, L. (2018). Improvement in scoliosis research society-22R pain scores after surgery for adolescent idiopathic scoliosis. Spine 43(*2*), 127–132. 10.1097/BRS.0000000000001978

Fernández-Rodríguez, R., Álvarez-Bueno, C., Cavero-Redondo, I., Torres-Costoso, A., Pozuelo-Carrascosa, D., Reina-Gutiérrez, S., Pascual-Morena, C., & Martínez-Vizcaíno, V. (2022). Best exercise options for reducing pain and disability in adults with chronic low back pain: Pilates, strength, core-based and mind-body. A network meta-analysis. The Journal of Orthopaedic and Sports Physical Therapy, 1–49. 10.2519/jospt.2022.10671

Gardner A, Cole A, Harding I. (2021). What does the SRS-22 outcome measure tell us about spinal deformity surgery for Adolescent Idiopathic Scoliosis in the UK? Ann R Coll Surg Engl., 103(*7*):530*-*535. doi: 10.1308/rcsann.2021.0005. PMID: 34192483; PMCID: PMC10335234.

Gremeaux, V., Casillas, J.M., Fabbro-Peray, P., Pelissier, J., Herisson, C., & Perennou, D. (2008). Analysis of low back pain in adults with scoliosis. Spine 33(*4*), 402–405. 10.1097/BRS.0b013e318163fa42

Grieder, S., Steiner, M.D. (2022). Algorithmic jingle jungle: A comparison of implementations of principal axis factoring and promax rotation in R and SPSS. Behav Res 54, 54–74. 10.3758/s13428-021-01581-x

Gür, G., Ayhan, C., & Yakut, Y. (2017). The effectiveness of core stabilization exercise in adolescent idiopathic scoliosis: A randomized controlled trial. Prosthetics and Orthotics International, 41(3), 303–310. 10.1177/0309364616664151

Hays, D. G., & Singh, A. A. (2023). Qualitative Research in Education and Social Sciences (2nd Edition). San Diego, CA: Cognella, Inc.

Jenkinson, C., Wright, L. & Coulter, A. (1994). Criterion validity and reliability of the SF-36 in a population sample. Qual Life Res 3, 7–12. 10.1007/BF00647843

Kerns RD, Turk DC, Rudy TE. The West Haven-Yale Multidimensional Pain Inventory (WHYMPI). Pain. 1985 Dec;23(4):345–356. doi: 10.1016/0304-3959(85)90004-1. PMID: 4088697.

Leopold, S. (2019). Editor’s spotlight/take 5: how common is back pain and what biopsychosocial factors are associated with back pain in patients with adolescent idiopathic scoliosis?. Clinical Orthopaedics and Related Research 477(4), 672–675. 10.1097/CORR.0000000000000689

Li, X., Shen, J., Liang, J., Zhou, X., Yang, Y., Wang, D., Wang, S., Wang, L., Wang, H., & Du, Q. (2021). Effect of core-based exercise in people with scoliosis: A systematic review and meta-analysis. Clinical Rehabilitation, 35(*5*), 669–680. 10.1177/0269215520975105

Marti, C. L., Glassman, S. D., Knott, P. T., Carreon, L. Y., & Hresko, M. T. (2015). Scoliosis Research Society members attitudes towards physical therapy and physiotherapeutic scoliosis specific exercises for adolescent idiopathic scoliosis. Scoliosis, 10(16). 10.1186/s13013-015-0041-z

Melzack R. The McGill Pain Questionnaire: major properties and scoring methods. Pain. 1975 Sep;1(3):277–299. doi: 10.1016/0304-3959(75)90044-5. PMID: 1235985.

Monticone, M., Ambrosini, E., Cazzaniga, D. et al. (2016). Adults with idiopathic scoliosis improve disability after motor and cognitive rehabilitation: results of a randomised controlled trial. Eur Spine J 25, 3120–3129. 10.1007/s00586-016-4528-y

Negrini, A., Negrini, M.G., Donzelli, S. et al. (2015). Scoliosis-Specific exercises can reduce the progression of severe curves in adult idiopathic scoliosis: a long-term cohort study. Scoliosis 10, 20. 10.1186/s13013-015-0044-9

Ng, S.-Y., Ho, T.-K., & Ng, Y.-L. (2019). Physical rehabilitation in the management of symptomatic adult scoliosis. IntechOpen. 10.5772/intechopen.81184

Nikolov, D. & Dimitrova, E. (2017). Correlation between pain and muscle strength in patients with adult scoliosis. Scripta Scientifica Salutis Publicae, 3(1), p. 33.

Papaioannou M, Diakomi M, Georgoudis G, Argyra E, Vadalouca A, Siafaka I. (2018). The Chronic Pain Grade Questionnaire: validity, reliability and responsiveness in Greek chronic hip pain sufferers. Hippokratia, (1):37–42. PMID: 31213756; PMCID: PMC6528698.

Physiopedia (2024). Oswestry disability index. https://www.physio-pedia.com/Oswestry_Disability_Index

Polaski AM, Phelps AL, Kostek MC, Szucs KA, Kolber BJ (2019). Exercise-Induced Hypoalgesia: A Meta-Analysis of Exercise Dosing for the Treatment of Chronic Pain. PLoS ONE 14(1):e0210418. 10.1371/journal.pone.0210418

Price DD, McGrath PA, Rafii A, Buckingham B. The validation of visual analogue scales as ratio scale measures for chronic and experimental pain. Pain. 1983 Sep;17(1):45–56. doi: 10.1016/0304-3959(83)90126-4. PMID: 6226917.

Reh, R., Mursidi, M.L., & Husin, N.A.A. (2011). Reliability analysis for pilot survey in i integrated survey management system. 2011 Malaysian Conference in Software Engineering, Johor Bahru, Malaysia, 2011, pp. 220–222, doi: 10.1109/MySEC.2011.6140673.

Roberts, H., & Dpt, H. R. (2023). Physical therapy management of hip pain in adults with scoliosis: A Case Series. Frontiers in Medical Case Reports. 10.47746/FMCR.2023.4202

Schreiber, J.B. (2021). Issues and recommendations for exploratory factor analysis and principal component analysis. Research in social and administrative pharmacy, 17(5), 1004–1011. 10.1016/j.sapharm.2020.07.027.

Schultz, L., Uyterhoeven, R., & Khalsa, S. (2011). Evaluation of a yoga program for back pain. Journal of Yoga & Physical Therapy, 1, 1–7. 10.4172/2157-7595.1000E103.

Schwab, F., Dubey, A., Pagala, M., Gamez, L., & Farcy, J. (2003). Adult scoliosis: a health assessment analysis by SF-36. Spine 28(*6*), 602–606. 10.1097/01.BRS.0000049924.94414.BB

Scispace. (2024). Why to use Exploratory factor analysis in pilot testing while developing scale? https://typeset.io/questions/why-to-use-exploratory-factor-analysis-in-pilot-testing-3oicomt9wl

Spark Chart. (2024). The crucial importance of conducting a survey pilot: unlocking the power of accurate data. https://www.sparkchart.com/conducting-a-survey-pilot/#:~:text=Piloting%20a%20survey%20helps%20ensure,the%20desired%20constructs%20or%20variables.

Steinmetz, L., Segreto, F., Varlotta, C., Grimes, K., Bakarania, P., Berdishevsky, H., Lanre-Amos, T. & Fischer, C.R. (2020). Surgeon attitudes toward physiotherapeutic scoliosis-specific exercises in adult patients with spinal deformities. International Journal of Spine Surgery, 13(6), 568–574. 10.14444/6079

Vasiliadis ES, Grivas TB, Kaspiris A. (2009). Historical overview of spinal deformities in ancient Greece. Scoliosis, 25;*4*:6. doi: 10.1186/1748-7161-4-6. PMID: 19243609; PMCID: PMC2654856.

Ware JE Jr, Sherbourne CD. The MOS 36-item short-form health survey (SF-36). I. Conceptual framework and item selection. Med Care. 1992 Jun;30(6):473–83. PMID: 1593914.

Weiss, H.-R., Moramarco, K., & Moramarco, M. (2016.). Scoliosis bracing and exercise for pain management in adults-a case report. J. Phys. Ther. Sci. 28:2404–2407.

Whitmarsh, Christine (2023). Pain Management Strategies in People with Chronic Pain. [Unpublished Research Proposal].

Wong, C.K.H., Cheung, P.W.H., Samartzis, D., Dip-Kei Luk, K., M. C. Cheung, K., Lam, C.L.K., Pui Yin Cheung, J. (2017). Dataset used to generate the results of this study.. PLOS ONE. Dataset. 10.1371/journal.pone.0175847.s001

Zaina, F., Marchese, R., Donzelli, S., Cordani, C., Pulici, C., McAviney, J., & Negrini, S. (2023). Current knowledge on the different characteristics of back pain in adults with and without scoliosis: a systematic review. Journal of Clinical Medicine, 12(*16*), 5182. 10.3390/jcm12165182

