## Appendices and Supplements for "Adult Scoliosis and Exercise: A Survey Instrument Pilot Study"

Christine Whitmarsh, MS

SUPPLEMENTAL MATERIALS

#### Appendix A: SE-18 rev 1.0

Q1 Your age now:

- ◯ 20-30 (1)
- ◯ 31-40 (2)
- ◯ 41-50 (3)
- ◯ 51-60 (4)
- ◯ 61-70 (5)
- ◯ Over 70 (6)

Q2 Gender assigned at birth:

- ◯ Female (1)
- ◯ Male (2)

Q3 Age when diagnosed with scoliosis:

○ Under 12 (1)

○ 12-16 (2)

○ 17-21 (3)

○ Over 21 (4)

Q4 Degree of spinal curvature:

○ 10-40 (1)

○ 41-60 (2)

○ 61 or greater (3)

Instructions:

For the following questions, you will be asked to describe your scoliosis pain, how you cope with your pain, and your exercise habits. Scoliosis pain is defined as any physical or mental pain that you know or believe to be related to the curvature of your spine. If you believe it to be related to your scoliosis, it is scoliosis pain. Take your time reading each question and following the instructions carefully when answering each item.

Q5 How long have you experienced scoliosis pain?

○ Since before being diagnosed. (1)

- Since being diagnosed. (2)
- 1-5 years after being diagnosed. (3)
- 5-10 years after being diagnosed. (4)
- More than 10 years after being diagnosed. (5)

Q6 How frequently do you experience scoliosis pain?

- Daily (1)
- Multiple instances per week (2)
- Multiple instances per month. (3)
- Less than one instance per month. (4)

Q7 Rate your overall scoliosis pain on a typical day (0 = no pain, 100 = worst pain):

| | 0 | 10 | 20 | 30 | 40 | 50 | 60 | 70 | 80 | 90 | 100 |
|---|---|---|---|---|---|---|---|---|---|---|---|
| When you wake up in the morning: () | | | | | | | | | | | |

| |
|---|
| By midday: () |
| By the end of the day: () |
| After standing for an hour or more: () |
| After sitting or laying in one position for an hour or more: () |

Q8 Rate your scoliosis pain in the following areas on a typical day (0 = no pain, 100 = worst pain):

0 10 20 30 40 50 60 70 80 90 100

| |
|---|
| Upper Back () |
| Middle Back () |
| Lower Back () |
| Hips () |

| |
|---|
| Arms () |
| Legs () |
| Neck () |
| Other: () |

Q9 Rate the intensity of these types of pain that you experience (0 = no pain like this, 100 = the worst of this type of pain):

| | 0 10 20 30 40 50 60 70 80 90 100 |
|---|---|
| Tightness () | |
| Sharp/Stabbing () | |
| Dull/Achy () | |
| Numbness/Tingling () | |
| Shooting/Radiating to Other Body Parts () | |

Q10 Which of these activities relieve your scoliosis pain? (Check all that apply.)

- ▢ Lying Down/Resting (1)
- ▢ Medication (2)
- ▢ Exercise (3)
- ▢ Stretching (4)
- ▢ Other: (7) ________________________________________________
- ▢ None of the Above (8)

Q11 Do you exercise?

- ◯ Yes (1)
- ◯ No (2)

Q12 If you exercise, how frequently?

- ◯ Daily (1)

○ Once a Week (2)

○ More Than Once a Week (3)

○ A Few Times a Month (4)

○ Rarely (5)

Q13 What kinds of exercises do you do? (Check all that apply.)

☐ Yoga (4)

☐ Pilates (5)

☐ Strength Training/Weights/Conditioning (6)

☐ Stretching (7)

☐ Other (8) ________________________________________________

Q14 Does exercise make your scoliosis pain:

○ Better (1)

○ The Same (2)

○ Worse (3)

Q15 How much does your pain restrict your physical movement? (0 = not at all, 100 = the most possible)

| | 0 10 20 30 40 50 60 70 80 90 100 |
|---|---|
| On a Typical Day () | |
| When Not Exercising Regularly () | |
| When Exercising Regularly () | |

Q16 How open are you to the idea of exercise as scoliosis pain relief?

- ○ Extremely (1)
- ○ Maybe (2)
- ○ Not At All (3)

Q17 Do you believe you would exercise regularly if it helped relieve your scoliosis pain?

- ○ Yes (1)
- ○ Probably (2)
- ○ Maybe (3)
- ○ No (4)

Q18 Which exercises would you consider doing if it would relieve your pain in the long-term?

- ☐ Machines (treadmill, elliptical, stationary bike, etc.) (1)
- ☐ Running/Jogging (2)
- ☐ Strength Training/Weights/Conditioning (3)
- ☐ Yoga (4)
- ☐ Pilates (5)
- ☐ Stretching (6)
- ☐ Other (7) ________________________________________________

### Appendix B: SE-18 rev 2.0 (in progress!)

Q1 Your age now:

- ○ 20-30 (1)
- ○ 31-40 (2)
- ○ 41-50 (3)
- ○ 51-60 (4)
- ○ 61-70 (5)
- ○ Over 70 (6)

Q2 Gender assigned at birth:

- ○ Female (1)
- ○ Male (2)

Q3 Age when diagnosed with scoliosis:

- ○ Under 12 (1)
- ○ 12-16 (2)

◯ 17-21 (3)

◯ Over 21 (4)

Q4 Degree of current spinal curvature:

◯ 10-40 (1)

◯ 41-60 (2)

◯ 61 or greater (3)

Instructions:

For the following questions, you will be asked to describe your scoliosis pain, how you cope with your pain, and your exercise habits. Scoliosis pain is defined as any physical or mental pain that you know or believe to be related to the curvature of your spine. If you believe it to be related to your scoliosis, it is scoliosis pain. Take your time reading each question and following the instructions carefully when answering each item.

Q5 If you have had surgery for your scoliosis, how much did the surgery improve your pain?

(VAS/slide style from “not at all” through “zero/no change’ to “a lot.”)

Q6 How long have you experienced scoliosis pain?

- ○ Since before being diagnosed. (1)
- ○ Since being diagnosed. (2)
- ○ 1-5 years after being diagnosed. (3)
- ○ 5-10 years after being diagnosed. (4)
- ○ More than 10 years after being diagnosed. (5)

Q7 How often do you experience scoliosis pain?

- ○ Daily (1)
- ○ Multiple times per week (2)
- ○ Multiple times per month. (3)
- ○ Less than once a month. (4)

Never

OR

VAS from never to daily?

Q8 Rate your overall scoliosis pain on a typical day (0 = no pain, 100 = worst pain):

| | 0 10 20 30 40 50 60 70 80 90 100 |
|---|---|
| When you wake up in the morning: () | |
| By midday: () | |
| By the end of the day: () | |
| After standing for an hour or more: () | |
| After sitting or laying in one position for an hour or more: () | |

Q9 Rate your scoliosis pain in the following areas on a typical day (0 = no pain, 100 = worst pain):

| | 0 10 20 30 40 50 60 70 80 90 100 |
|---|---|
| Upper Back () | |

| |
|---|
| Middle Back () |
| Lower Back () |
| Hips () |
| Arms () |
| Legs () |
| Neck () |
| Other: () |

Q10 Rate the intensity of these types of pain that you experience (0 = no pain like this, 100 = the worst of this type of pain):

0 10 20 30 40 50 60 70 80 90 100

| |
|---|
| Tightness () |
| Sharp/Stabbing () |

| |
|---|
| Dull/Achy () |
| Numbness/Tingling () |
| Shooting/Radiating to Other Body Parts () |

Q11 Which of these activities relieve your scoliosis pain? (Check all that apply.)

Arrange the following in order from the activities that relieve your scoliosis pain the BEST to those that relieve it the LEAST.

- ▢ Lying Down/Resting (1)
- ▢ Medication (2)
- ▢ Exercise (3)
- ▢ Stretching (4)

-Add more based on frequent open ended fill ins from 1.0

Q12 How often do you exercise?

- Daily (1)
- Once a Week (2)
- More Than Once a Week (3)
- A Few Times a Month (4)
- Rarely (5)

Never

Q13 What kinds of exercises do you do? (Check all that apply.)

- Yoga (4)
- Pilates (5)
- Strength Training/Weights/Conditioning (6)
- Stretching (7)

-Add in frequent fill ins + PSSE

Q14 How does exercise affect your scoliosis pain?

VAS - makes it worse > makes it better, zero is unchanged

Q15 How much does your pain restrict your physical movement? (0 = not at all, 100 = the most possible)

| | 0 10 20 30 40 50 60 70 80 90 100 |
|---|---|
| On a Typical Day () | |
| When Not Exercising Regularly () | |
| When Exercising Regularly () | |

### Supplement C: Planned Mixed Methods Study

**Research Proposal:**

***Adult Scoliosis and Exercise, a Mixed Methods Study***

#### Research Methods and Design

##### Study Overview with Classification

##### Research Questions

1) Quantitative: What is the effect of a 6-month regimen of scoliosis-specific exercises on adults with chronic scoliosis pain?

2) Qualitative: How do scoliosis-specific exercises affect individual perceptions of day-to-day pain and mobility in adults with chronic scoliosis pain?

##### Sampling Plan

The following list of sampling options (Martella, et al., 2013) could potentially be applicable to this study.

- Nonprobability sampling: This sampling method would be applicable since it would be impossible to determine the size of the complete population of adults with scoliosis who experience pain and exercise even occasionally. With nonprobability sampling, there is an unknown likelihood of any single individual being selected from the population. This method usually generates samples that are not necessarily representative of the full population (as described above). The following are types of nonprobability sampling.

- Convenience sampling: Samples participants who are available to the researcher; difficult to generalize results.
- Opportunity sampling: Samples participants who are willing and available.
- Volunteer sampling: Samples participants who self-select based on study recruiting (ex. from the SE-18 survey instrument - a social media graphic with QR code leading them to survey).
- Snowball sampling: Using the existing group of participants (e.g. through the methods above in this case) to recruit additional ones from their network (ex. from SE-18 - a physical therapist friend who works exclusively with scoliosis patients, emailed survey info to her mailing list).
- Purposive sampling: Hand picking participants that are likely to produce quality data for the study.

For this mixed methods study, a combination of nonprobability sampling methods listed above will likely be utilized. Of note is that nonprobabilistic sampling does not necessarily mean that a study is without meaning or value, but rather that the study results will be less generalizable than when probabilistic sampling methods are used (Martella, et al., 2013).

**Study Variables and Measures**

**Quantitative Variables (RQ1)**

RQ1 (Quantitative): What is the effect of a 6-month regimen of scoliosis-specific exercises on adults with chronic scoliosis pain?

A review of the literature shows at least one longevity recommendation of 3-9 weeks (Fernández-Rodríguez, et al., 2022) and a much longer one with a duration of 35 months (Gur, et al., 2017). 6 months is a tentative recommendation, subject to change based on planned consultation with a specialist in this area.

The independent variables assigned to help answer this question, based on the review of literature, will be (regimens of) Pilates, PSSE exercises, traditional physical therapy, and core strengthening/stability exercises.

The conceptual variable being measured here, is the theoretical relationship between scoliosis pain and exercise. After researching several different applicable scales, it was decided that the dependent variables will be measurements from the following 3 scales: the SRS-22, a scoliosis-specific pain, mobility, and quality of life scale, the VAS scale, a standard measurement of pain severity, and, optimistically, this researcher's SE-18 scale.

**Qualitative Variables (RQ2)**

RQ2: How do scoliosis-specific exercises affect individual perceptions of day-to-day pain and mobility in adults with chronic scoliosis pain?

Virtual focus groups will be utilized to measure this RQ, due to the researcher-observed effectiveness of scoliosis peers discussing their condition within the chronic pain and scoliosis communities. Rich descriptions will be helpful to understand the quantitative measurements generated from RQ1.

The overall conceptual variable in evaluating this question, is the perception of how exercise impacts participants' day-to-day scoliosis pain and mobility. Operational variables might include inter-observer agreement of coding (coefficients), outsider agreement of the researcher interpretation of the qualitative data, triangulation of data. Confounding variables could be similar to confounding variables in RQ1, centered on participants' unique perceptions of the impact of scoliosis on their body, in pain, disability, mobility and quality of life and their compliance with and attitudes toward the exercise interventions.

**Measures - RQ1**

A MANOVA will most likely be utilized for statistical measurement of quantitative data from RQ1. However, Canonical Correlation Analysis (CCA) is also an option as it could offer multiple viewpoints on how different types of exercises are related to different scale measurements (ex. Pilates-SRS-22, PSSE-SE-18, and such).

The advantage of MANOVA for a study like this would be in creating collective assessments, as in, measuring if the type of exercise has a significant impact on the collective scale measurements (SRS-22, VAS, SE-18) while also, through post hoc ANOVA testing, identifying significant relationships between types of exercises and individual scales through pairwise comparisons (Martella, et al., 2013).

MANOVA also offers increased control over Type 1 error rates without sacrificing statistical power, thus making it a good choice for analysis of complex data like in this proposed study (Spector, 1981). Finally, when applied specifically to healthcare data, according to Statistics Solutions (2024), MANOVA offers the benefit of revealing how, in the case of this research, different types of exercise simultaneously influence all 3 pain scale measurements, rather than looking at the impact of the IVs on each scale by itself.

A drawback of MANOVA that might require an alternate course of action would be its assumptions:

1) Normal distribution of data.
2) Equal group variances in dependent variables.
3) Independence of observations.

If assumptions are not met, a Multivariate Kruskal-Wallis (MKW) will be run instead.

In terms of measures related to the qualitative oriented RQ2, the plan is to measure the data, ultimately printed transcripts, resulting from focus groups and/or group interviews using a thorough coding process, using multiple coding methods. More details related to these procedures will be covered in the following section.

**Increasing Power**

The main power challenge with a study like this one lies in sample size. On one hand, the sample size should be robust enough to support the multivariate data analysis proposed to answer RQ1. On the other hand, the sample needs to be manageable enough for the qualitative piece of this mixed methods study and if a multiple baseline vs. QE design is utilized, that will also support the goal of a manageable sample size.

That being said, here are 5 methods (Martella, et al., 2013) of increasing power in a research study along with ideas to apply each one to this study.

**1) Using parametric vs. nonparametric tests.** Since this is largely dependent on how well this study's data, obtained from participants using a nonrandom, non-probability sampling method as previously detailed, will meet parametric assumptions

**2) Decrease sources of error variance.** This one might be more controllable, by running the most rigorous study, with the greatest amount of experimental control possible, and eliminating as many validity threats as possible, within the constraints of the chosen design(s).

**3) Relax the alpha level (ex. $p<.10$ vs. $p<.05$).** This can likely be applied to the MANOVA testing.

**4) One tailed rather than two tailed hypothesis, which decreases the possibility of Type I error.** This one is not applicable since the proposed research design does not involve a hypothesis. However, as noted in the previous section, utilizing the MANOVA method of data analysis decreases the possibility of Type I errors.

**5) Increase n#.** As previously noted - this is likely not possible..

#### Procedures

**RQ1: What is the effect of a 6-month regimen of scoliosis-specific exercises on adults with chronic scoliosis pain?**

For this first, quantitative phase of the mixed methods study, one thing observed in the literature review is that in the sparse experimental and quasi-experimental studies done in this subject matter area, researchers typically assign participants to different types of exercise - ex. Pilates vs. PSSE. Based on the research done as well as the researcher's deep subjective understanding of how scoliosis pain responds to various types of exercise, there seems to be new opportunities through a different approach. Since the objective is to collect quantitative data measuring differences in pain levels and mobility based on various types of exercises (IVs), the goal will be to rotate the entire group of study participants in increments through all of the exercise conditions.

The reasoning behind this procedure is the highly individualized nature of scoliosis pain. This is not necessarily attached to degree of curvature, whether spinal fusion has occurred, and time elapsed since diagnosis (hence the frustration of researchers trying to treat scoliosis pain). One participant could respond completely differently to pilates than the next person, and find better pain relief with PSSE, and so on.

Therefore, in order to rotate an entire group of participants through multiple treatment conditions, a multiple baseline design across behaviors might be an appropriate design for the quantitative piece of the study. Such a design might look as follows: create cohorts of participants, take baseline measurements, apply exercise A to the first group, stagger and do the same with the next cohort, and so on.

The question to be answered before a final design is selected is: How can cohorts of participants cycle through several different exercise regimens over the assigned time period, with measurements taken, using the 3 dependent variable measurement scales, measuring how each exercise impacts their pain and mobility levels? The main concern in using multiple baseline design is the built-in lack of external validity, generalization.

Data collection in this design will be self reported from the participants including an online journal (mainly for the qualitative piece). The subjects will undergo exercise interventions (specified by frequency, dosage, and intensity) with expert consultation and indirect supervision by: a) an exercise physiotherapist and/or physical therapist specializing in scoliosis treatment (ex. A Schroth treatment provider), and b) sign-off by an board certified orthopedist/ortho surgeon specializing in scoliosis.

Participant cohorts will receive an initial live virtual group training by the aforementioned exercise physiotherapist or PT at the beginning of each exercise intervention including a virtual Q&A session at the end of each session. Pre-recorded videos (form checks, motivation, different modifications, etc.) will also be made available to participants at designated points in the treatment intervention including a “you did it!” video at the end of the study with strategies for deciding which exercises worked the best for their body and how to continue their “new exercise habit” over the long term.

For the quantitative piece, an automated system or API, could be utilized, where participants can easily log the nuts and bolts of their weekly exercise data. An easy-to-use automated system would have the triple benefit of a) making it easy for the participants b) ease of data collection for this study and c) a tool that could be made available for researchers wishing to do replications.

**RQ2: How do scoliosis-specific exercises affect individual perceptions of day-to-day pain and mobility in adults with chronic scoliosis pain?**

Following the exercise intervention, once all cohorts have been cycled through all types of exercise, participants will be randomly assigned to 2-3 (due to typical scheduling issues) separate virtual focus groups. Time gaps will be utilized as cohorts are cycled through via additional pre-recorded exercise bonus videos, journaling exercises, videos on developing an exercise habit, and such provided to the earlier cohorts.

The objectives of the focus groups will be to: a) allow participants to share their experiences from the exercise program with one another, b) gain as much rich descriptive data as possible to answer RQ2 with compelling open-ended questions and probes (via standardized open-ended interview method) which will be made available as PDFs in the study for replication purposes, and c) gather metadata on the study itself for future fine tuning of replications. Focus groups will be recorded, transcribed, and thoroughly coded, with interobserver agreement done to ensure accuracy of coding.

**Missing Data**

Missing data will be handled using the most appropriate of the following missing data procedures. Although based on the SE-18 survey instrument pilot study and the missing data patterns observed, if a prediction needed to be made, based on the study - it would be MCAR/imputation.

1) MCAR means "missing completely at random." The probability of what is missing (IV, DV) is uncorrelated with the data. In other words, the data is missing totally by chance.

2) MNAR, "missing not at random," means that pieces of data are missing because of something the researcher has not measured.  In other words, the data is missing because of something we have not measured.

3) MAR, "missing at random," means the data is missing conditionally (as the instructor clarified) at random, meaning pieces of data are missing because of a specific IV that we HAVE

measured. In other words, we know why the data is missing and have measured that reason (and can therefore control it).

4) Multiple imputation is the "multiples" version of Stochastic Regression Imputation, wherein missing values are predicted from observed values, but error is added in. It is done multiple times to adequately increase standard errors resulting in a new regression model with more accurate predictions (than Stochastic or simple regression imputation). This accounting for standard error is why multiple imputation is a better choice than listwise deletion/deleting subjects.

**Anticipating Validity Considerations**

**Quantitative Validity**

Starting with the quantitative RQ1, assessing validity threats of both possible designs - QE/counterbalanced and multiple baseline.

| **QE/Counterbalanced Design** | | |
|---|---|---|
| **Internal Validity Threats Controlled** | Maturation, selection, statistical regression, mortality, instrumentation, testing effects, history. (because there is no non-intervention control group) | |
| **Internal Validity Threats Not Controlled & My Mitigation Strategies** | Threats related to lack of random assignment and due to my sampling method, lack of random selection:<br><br>1. Selection bias *(despite assurances to the contrary by our textbook; which also continued to reinforce that assessing validity threats should be done on a study-by-study basis - what I'm doing here!)*. A potential threat since I am using non probability sampling. | 1. First, there is the awareness (increased vigilance) of the inherent selection bias risks in both the sampling method and design selected. From there, and also as noted earlier in this proposal in the sampling method section, a strategy will be designed to put together the most diverse sample possible.<br><br>2. Utilizing the mitigation strategy in #3 below.<br><br>3. Mitigating by creating tools for |

| | | |
|---|---|---|
| | 2. History - ditto note above; a potential threat for mine since participants will be left to their own lives/routines and self apply the exercise treatment condition.<br><br>3. Mortality/Attrition: same as above, going with the case-by-case and seeing this as a potential sizable threat in my study, since the treatment condition is self-directed exercise.<br><br>4. Multiple treatment interference: Calling this… possibly. If participants have an existing exercise routine in place.<br><br>5. Social desirability bias: Another possibility, under the phenomenon of people who never exercise but then when they finally hit the gym they post it on social media to get "social desirability"points from their network.<br><br>6. Order effect: Any potential issues arising from the order of exercise interventions. | participants to stay engaged with exercise regimens (journaling, app/system for checking in, motivational/strategy premade videos and such).<br>4. Prescreening of participants as thoroughly as possible.<br><br>5. Preventing direct contact with researcher during study to prevent favoritism and other similar biases.<br><br>6. Doing careful research including with the physiotherapist/PT expert(s) to design the most optimal order of exercise interventions. |
| **External Validity Threats Controlled** | Novelty and disruption, Hawthorne, experimenter - because of this design's ongoing implementation of different IVs (exercise methods). | |
| **External Validity Threats Not Controlled & My Mitigation Strategy** | Population Validity/Generalizability (no random assignment nor random selection in this design). | Similar to above - awareness and in this case, full disclosure in the writing up of the final study, as seen in multiple such studies throughout this semester. Clearly communicating the strengths and limitations of the study AND encouraging replications, in the write up so CRCs can make informed decisions about what they ethically should and should not use the results for in practice. |

| Multiple Baseline Designs - Internal Validity Issues | | | | As Applied to This Study |
|---|---|---|---|---|
| **Condition Length** | 3+ stable data points or a counter therapeutic trend | # of data points increases as # of baselines increases | | Will assess at time of study. |
| **Independence of Behaviors** | *Critical rule. | Changes in one behavior shouldn't coincide w/changes in others. (Like seen in multiple baseline design across participants.) | Textbook states that diffusion of treatment might be an issue depending on the researcher but in their table it says controlled? | So, for example, changes in Pilates effects of cohort A should not coincide with changes in core conditioning effects of cohort B. |
| **Amount of Data Overlap** | Need 3+ overlapping data points for 1st baseline, 3+ for 2nd, 3+ during behavior #1 intervention. Total of 6+ data points for 2nd baseline. | Experimental control is because it shows that prior person's behavior would have stayed the same without intervention. Other person's behavior stays stable. | CRC - look at amount of data overlap + trend. | Noted. |
| **Number of Baselines** | Need 2+<br><br>But are 2 enough to show experimental control? (see below) | | | Noted. |
| **Level and Rapidity of Change** | *Critical. | Changes in data need to be significant and rapid in each of the behaviors. | More support for - better to have more than 2 tiers (or else have an A-B, weaker design). | Noted. |
| **Withdrawal** | Increases internal validity in m-b designs by withdrawing the | Implementation of treatment is staggered across multiple baselines, | | Noted. |

| | | |
|---|---|---|
| | treatment condition from each behavior, participant, or setting. | but each baseline has a withdrawal condition. |

| Multiple Baseline Designs - External Validity Issues | |
|---|---|
| Generalization to people (population) and other situations (ecological). | Withdrawal design lacks external validity. |
| Best to determine generalization/ext validity through replications across behaviors, participants, and/or settings vs. through a single M-B design. | Primary reason for MB design is to control for INTERNAL not external validity. |
| M-B designs, alt. to withdrawal designs, have stronger external validity, more versatile, and are better reflective of real world | |

**Qualitative Validity**

Finally, addressing the various validity threats for the qualitative piece of this mixed methods study. Applying the concept of validity to qualitative designs is largely based around an outsider's assessment of the apparent truthfulness of the data collected. Therefore, qualitative validity is not associated with design (as it is in quantitative designs) but rather with the authenticity and believability of the data collected. As a result, concepts of external validity like population and ecological validity and overall generalizability translate to an overall recommendation for the CRC to use a qualitative study as fuel for future replications. However, in the interest of ensuring all future study bases are covered,  following is a more detailed breakdown.

**Managing Validity Threats in Qualitative Studies**

| | Defined As | Key Point(s) | CRC Concerns | As Applied To This Study |
|---|---|---|---|---|
| **Descriptive Validity** | Accuracy of data in representing phenomenon of interest. | Assessed in terms of what was included and/or excluded from researcher's account. | What was included/excluded, agreement from outsiders. | Document, document, document! |
| **Interpretive Validity** | Subjective meaning of objects, events, behaviors; that which cannot be observed. | Researcher's job is to construct participant meaning through observation of words and behaviors. | Researcher method to gain this understanding necessary to make valid interpretations. | Diligent attention to construction of focus group questions, and coding process, including via inter observer agreement. |
| **Theoretical Validity** | "Maintaining integrity to ensure accurate interpretation of data." | Explanation + description + interpretation of subject account. Community's interpretation and acceptance of this account. | How was theory constructed, do others agree that it's solid, are descriptions and interpretations accurate and how were concepts from data put together. | Infusing as much theory from frequent updated reviews of the literature and researcher's own work from different angles to garner peer validity. |
| **Generalizability** | Ability to extend study's results to other people, settings, times. | Similar to quant ext val. But more logical or subjective.<br><br>Internal: ability to generalize within group being studied now to those outside the group.<br><br>External: Ability to extend results to outside communities and | Does theory make sense for the subjects (and stakeholders?) and will it hold up in other situations, conditions, groups of people? Be aware of type of gen. attempted and determine if theory makes sense for other situations/conditions. | The goal here is to do multiple research studies, each one providing more info than the last to feed the next one AND each one with better experimental control/rigor to continue building a case for generalizability of research.<br><br>*A personal note from* |

| | | | | |
|---|---|---|---|---|
| | | groups. | | *researcher: I'm not here to only help my study participants manage their scoliosis pain and provide yet more "vague" data for scoliosis clinicians to discount or disregard (like the ample case studies). I want to help as many people like me, as humanly possible. Now, thanks to this class, I understand the level of experimental control that will be required to accomplish this goal.* |
| **Evaluative** | Making valid statements. | How well researcher evaluation matches public evaluation of findings. | | |

**Anticipated Results/Findings – What results are expected given the design?**

Given the current status of this long-term research mission and the combination of methods selected for the design described in this proposal, the anticipated results of this study can best be categorized as - targeted findings about which types of exercises have the greatest overall impact on participants' scoliosis pain, mobility, and overall quality of life.

The results from this mixed method study, given its design constraints including a lack of control group, random selection of participants, and random assignment, while informative and hopefully fuel for future replications, will likely not be generalizable (as a standalone body of evidence) to larger populations of adults with scoliosis. But for where this particular study is situated within the greater mission - it is anticipated to serve its unique purpose. Following, is a more detailed breakdown of anticipated results for each aspect of the overall mixed method design.

**Quantitative**

**QE/Counterbalanced Design**

Utilizing this type of design for the quantitative piece, answering RQ1, would offer better experimental control (due to QE aspect) and, based on the elements of the counterbalanced design, a more rigorous examination of how the multiple independent variables (types of exercise) impact various pain scale measurements. This design, as it turns out, might be best positioned to answer RQ1 (time will tell): What is the effect of a 6-month regimen of scoliosis-specific exercises on adults with chronic scoliosis pain?

It is anticipated that the quantitative scale data gathered using this design would also help to fine tune the exercise items in the newly minted SE-18 scoliosis pain and exercise scale. As noted previously in this proposal, additional detail is required, regarding which exercises have which impact on pain/mobility for adults with scoliosis.

*Or*

**Multiple Baseline Design**

With a lesser amount of experimental control than the QE/counterbalanced design, it is anticipated that the findings from this type of design would center more on the individual differences (by cohort) between baseline and individual types of exercise treatments.

**Data Analysis via MANOVA**

MANOVA is testing the impact of predictor/treatment variables, in this case, types of exercise, on combined outcome variables, here, 3 different pain/mobility assessment scales. For the MANOVA to be significant, there must be a difference between treatments for one or more of the outcomes (scales). It is predicted that significant differences will be found between types of exercises based on their measurements on the 3 pain scales.

At that point, assuming significant differences are found, additional tests, including for effect sizes and a discriminant analysis, will be run in order to obtain more information about the difference in impact of the independent variables on the dependent variables (i.e. which types of exercises impacted which scales and how). Here, we come to possibly the most valuable part of the study - which type of exercise(s) has the greatest impact on the pain/mobility/QOL of adults with scoliosis? Pilates, PSSE, or core? Then, future replications using other types of exercise, with different frequencies of exercise and over different time periods, can be run to continue this work.

**Qualitative**

In regards to the answering of RQ2, “How do scoliosis-specific exercises affect individual perceptions of day-to-day pain and mobility in adults with chronic scoliosis,” anticipated results are reflective the broad range of results and responses seen in past research studies and in the SE-18 pilot study, about which types of exercise work best as scoliosis pain management strategies.

With a focus group atmosphere, it is expected that a similar range of responses, detail, and personalizations to each participant, will also be seen. From the outside, this might seem like a negative - yet more qualitative, one off data. But in this case, in the context of this mixed methods approach, the results of the qualitative piece of this study could be effective in explaining (via the planned sequential explanatory strategy) and expanding on the data obtained in the quantitative portion of the study.

**Conclusion**

In the way of context, here is the research journey leading up to the study detailed in this proposal with takeaways and how each step helped determine the next.

1. A small phenomenological study (n=3) on chronic pain and exercise.
    a. Major takeaways: Chronic pain is highly individualized and of the 3 participants, the one that exercised regularly had the best control of their pain, the one that occasionally exercised had some pain control, and the participant that did not exercise at all was nearly incapacitated from chronic pain.
    b. Next research steps inspired: A second, more comprehensive coding process to gain the deepest possible understanding of chronic pain.
2. The decision to create a scoliosis pain specific & exercise survey instrument - literature review.
    a. Major takeaways: As noted earlier in this proposal, the literature review showed very few research studies on this subject area with designs featuring good

experimental control (ex. RCTs). Also noted was the existence of one predominant survey instrument to measure scoliosis pain, but with no real representation of exercise in the scale.

   b. Next research steps inspired: Develop a SE-18 pilot study to fine tune an eventual scale to study the relationship between scoliosis pain and exercise.

3. SE-18 pilot study
   a. Major takeaways: More information is needed on the exercise piece of the relationship to better fine tune the SE-18 for more widespread use with better reliability.
   b. Next research steps inspired: Gain a better understanding of which types of exercise have the most impact on multiple pain/mobility/QOL scales.
4. The mixed methods study outlined in this proposal.
   a. Major takeaways predicted: Quantitative findings on which exercises most impact pain/mobility/QOL levels in adults with scoliosis, accompanied by qualitative detail to further understand the relationship, all for the purpose of improving the SE-18 for another, larger rollout.
   b. Next research steps: Use what has been learned to create the SE-18 rev 2.0. After that, ideally, utilizing the SE-18 as part of an even larger QE utilizing the most experimental control possible, for the most generalizable results possible.

In relation to the prior work outlined above, the mixed methods research design outlined in this proposal appears to be the appropriate next research step in this long-term journey. It is unclear what further predictions can be made at this time based on the available information.
